# Evaluating the cost-effectiveness of rapid diagnostic testing for the identification of pathogens and resistance genes in bloodstream infections

**DOI:** 10.1101/2025.06.12.25329483

**Authors:** James K. Karichu, Mark Pennington, Xiyu Bao, Kiera Lander, Tiffeny T. Smith, Adam Thornberg

## Abstract

**Objectives:** Bloodstream infections (BSI) are a leading cause of mortality worldwide. Rapid detection of the causative pathogen can help optimise therapy, reduce mortality, curb antimicrobial resistance, and lower healthcare costs. This study evaluates the cost-effectiveness of adding molecular rapid diagnostic tests (mRDTs) to microbiology standard-of-care (SoC) methods. mRDTs evaluated include the Cobas^®^ Eplex blood culture identification (BCID) panels, BioFire^®^ BCID panel, BioFire^®^ BCID2 panel, Accelerate PhenoTest^™^ blood culture (BC) kit and Diasorin Verigene^®^ BCID panels.

**Methods:** A decision-tree model was built to quantify the incremental costs and outcomes associated with adding mRDTs to the SoC. The inputs were derived from the published literature. The analysis considered a population aged 65 years and 45% female, admitted to a United States (US) hospital with a suspected BSI. Model outcomes included costs, 30-day mortality, quality-adjusted life years (QALYs) and adverse events (*Clostridioides difficile* infection and acute kidney injury [AKI]). A United Kingdom (UK) setting in place of the US setting was also considered in the scenario analysis.

**Results:** A strategy involving the Cobas Eplex BCID panels as an adjunct test to the SoC dominated SoC alone without Cobas Eplex BCID panels, saving $164 per patient and averting 24 deaths per 10,000 patients. Earlier optimisation of ineffective empiric therapy generated half of the lives saved, with the majority of the remainder from reductions in AKI. This strategy was also dominant compared with other mRDTs. In a UK setting, Cobas Eplex BCID panels remained cost-effective, saving £51 compared with SoC. Results were robust to scenarios varying key model inputs including time to pathogen identification with SoC.

**Conclusions:** The model demonstrated improved patient survival and reduced average total costs with mRDT. The Cobas Eplex BCID panels, which has the largest pathogen coverage, reduced both mortality and overall costs compared with other mRDTs.

**Key points for decision makers:**

- Rapid identification of BSI pathogens enables early optimisation of antimicrobial therapy, which can improve patient outcomes, reduce healthcare costs, and mitigate antimicrobial resistance. Conventional methods, including culture followed by MALDI-ToF MS, are time intensive. In contrast, mRDTs can identify pathogens within hours, facilitating earlier treatment optimisation.
- Evidence on the cost-effectiveness of mRDTs is limited, despite their potential to reduce overall costs by reducing length of stay (LOS). This study provides a comprehensive assessment of the cost-effectiveness of adding mRDTs to SoC.
- Cost savings from reductions in LOS and adverse events arising from broad-spectrum antimicrobial therapy are more than sufficient to offset the cost of procuring the mRDTs. Hence the use of mRDTs improves patient outcomes and contributes to antibiotic stewardship, alongside lowering costs.
- The Cobas Eplex BCID panels, with broadest coverage of pathogens, dominated other mRDTs.

**Plain Language Summary:** Bloodstream infections (BSI) are responsible for significant morbidity and mortality, causing 250,000 deaths each year in Europe and the United States (US) [1]. Rapid treatment with an effective antimicrobial is potentially one of the most important elements of care for patients with BSI, and patients are typically treated with two broad-spectrum antimicrobials rapidly (within an hour) if BSI is suspected. Conventional methods to identify the cause of the BSI require culturation of blood culture bottles followed by Gram stain and sub-culture onto solid agar plates to isolate individual colonies. Molecular rapid diagnostic tests (mRDTs) allow detection of a range of different pathogens causing BSIs within a few hours of a blood culture bottle flagging positive [2]. These tests offer the potential to improve care by reducing time to appropriate therapy which in turn can save lives and lower costs. This analysis used a decision-tree model to evaluate the costs and benefits of adding mRDTs to conventional methods. The study finds that mRDTs save lives and generate lower overall costs compared with standard of care (SoC) methods alone. Among the mRDTs, the Cobas^®^ Eplex BCID panels, which detect the broadest spectrum of pathogens, were associated with the highest reduction in mortality and overall costs compared with other mRDTs.

**Declarations:** *I. Funding:* COBAS and EPLEX are trademarks of Roche. All other product names and trademarks are the property of their respective owners. Financial support for this research analysis was provided by GenMark Diagnostics (A member of the Roche Group) to Source Health Economics—content experts (MP, XB, KL), and medical writing support was provided by Dr. Dom Partridge of Source Health Economics.

*II. Conflicts of Interest:* JKK and AT are employees of Roche Molecular Systems (RMS). JKK and AT hold Roche stock options. TS is an employee of Roche Diagnostics Corporation (RDC).

*III. Availability of data and material:* The analysis used secondary data sources which are publicly available. No primary individual patient data was collected or used.

*IV. Ethics approval:* The analysis used secondary data sources. No individual patient data was collected or used. Consequently, ethical approval for the study was not required.

*V. Consent to participate:* No individual patient data was collected or used, and hence consent was not required.

*VI. Code availability:* The economic model is considered commercially sensitive and we are unable to provide access to it.

*VII. Author contributions:* JKK and AT conceived the study. MP, JKK, AT and KL designed the study. MP, JKK, AT, TS and XB compiled the data. MP, JKK, AT, TS, KL and XB reviewed the results. MP drafted the manuscript. MP, JKK, AT, TS, KL and XB reviewed the manuscript.

## 1 Introduction

Bloodstream infection (BSI) generally refers to the presence of pathogenic microorganisms in the blood, often causing systemic inflammatory responses and potentially leading to severe conditions such as sepsis [3–5]. If BSIs are not treated quickly and effectively, they can lead to sepsis-induced organ dysfunction [6]. In the most severe cases, sepsis may progress to septic shock, and severe organ dysfunction including respiratory failure, heart failure, kidney failure, and death. Despite advances in treatment, BSI remains a major cause of morbidity and mortality worldwide [7, 8]. The estimated burden of BSI in Europe and North America is 2,000,000 episodes each year, of which 250,000 result in death [1].

Prior to definitive determination of the causative pathogen(s), rapid empirical treatment is required to minimise the risk of morbidity and mortality [9, 10]. Although rapid treatment with broad-spectrum antibiotics is associated with risks (e.g. *Clostridioides difficile* infection and acute kidney injury [AKI]), delayed diagnosis and initiation of appropriate treatment is associated with poorer outcomes [11]; with infections potentially leading to increased long-term disability, excess length of stay (LOS) in hospitals, large additional costs for health systems, as well as loss of quality of life (QoL) for patients and their families [12].

While practice varies across laboratories, standard of care (SoC) testing methods for diagnosis of BSIs generally combine matrix-assisted laser desorption ionization–time of flight mass spectrometry (MALDI-ToF MS) with conventional culture techniques and biochemical tests. Whilst MALDI-ToF MS can expedite pathogen identification (ID), especially in scenarios where a laboratory is using a direct from blood culture ID protocol, subculturing is still required to obtain pure colonies, and this step prolongs the time to pathogen ID, while also not providing any information on whether the pathogen is resistant or susceptible to any antibiotics. Recently, conventional culturing approaches including MALDI-ToF MS have been complemented by molecular rapid diagnostic tests (mRDT) to speed up diagnosis and improve accuracy [2]. These tests generally use: *in situ* hybridization-based methods (A); Deoxyribonucleic acid (DNA) micro-array based methods (B); or nucleic acid amplification-based methods (C) [13]. The most commonly used mRDTs include Accelerate PhenoTest^™^ BC (method A), Diasorin Verigene^®^ Gram-Positive and Gram-Negative Blood Culture Tests (method B), BioFire^®^ Blood Culture Identification (BCID) and BioFire^®^ BCID2 Panels (method C), and Cobas^®^ Eplex BCID panels (method A and C) [14]. Of the molecular methods, all of these platforms and their respective panels are Food and Drug Administration (FDA) cleared and capable of detecting a range of Gram-positive and Gram-negative bacteria. All except Accelerate PhenoTest can detect common resistance mechanisms such as KPC, VIM, OXA, *mecA, mecC, vanA*, and *vanB*, among others, and all except Diasorin Verigene BCID panels can detect fungal pathogens.

While mRDTs may increase direct procurement costs for the clinical microbiology laboratory, they can lower total hospital costs and improve patient outcomes through prompt optimisation of therapy following ID of organisms and their resistance markers. mRDTs have the potential to save money through, for instance, reductions in hospital LOS [15]. Published economic evaluations have indicated that mRDTs are generally cost-effective but are subject to a number of limitations [16, 17]. All have used a decision-tree format and applied payoffs for LOS and mortality as a function of the use of (a specific type of) mRDT or, the ability of the mRDTs to detect the organism. This approach has limitations, as it does not accurately capture differences in time to effective therapy for resistant strains, does not reflect the benefits of earlier transfer to targeted therapy, and does not reflect differences in mortality (or LOS) across different types of pathogens causing BSI. These limitations are magnified when trying to compare one mRDT with another. This analysis seeks to address these limitations by evaluating the incremental costs and benefits of adding mRDTs to SoC methods.

## 2 Methods

### 2.1 Model overview

The model considers the cost-effectiveness of mRDTs as an adjunct to the SoC. SoC is defined as conventional culture and MALDI-ToF MS without mRDT. SoC may require 16–24 hours (post Gram stain) to generate pathogen ID results and can be variable. The analysis population consists of hospitalised adults suspected of BSI. The primary country setting is the United States (US), with the United Kingdom (UK) considered in a separate scenario. The perspective of the analysis is a US hospital, whereas in the UK scenario, the perspective is the National Health Service (NHS). The primary outcomes are incremental costs and the quality-adjusted life years (QALYs) gained per patient. Additionally, the analysis estimates deaths averted and adverse events (*C. difficile* infection and AKI). A lifetime horizon is applied to the analysis in order to estimate QALYs gained. A 30-day time horizon is applied to costs and other intermediate outcomes in the model e.g., *C. difficile* and AKI cases averted. Discounting was applied to QALYs at 3% per year (3.5% for UK scenario) following recommended guidance [18, 19]. Costs were not discounted given the short time horizon for resource use (30 days).

The model was designed following consideration of the clinical pathway for a patient admitted to hospital with a suspected BSI and the key points at which test results might influence outcomes (see supplementary material section S1.1.). Guidelines recommend that patients with a suspected BSI be rapidly started on broad-spectrum antimicrobial therapy [20, 21]. Treatment may be optimised following pathogen ID (and resistance determination, if applicable). If the broad-spectrum therapy is ineffective, it could be changed to an effective targeted therapy. If the broad-spectrum therapy is effective, optimisation (i.e. de-escalation) to a targeted therapy which is also effective but associated with a lower risk of adverse events, may be considered. Finally, treatment may be further modified if the antimicrobial susceptibility test (AST) or antimicrobial resistance genetic results suggest presence of a resistance mechanism that renders the current therapy ineffective or susceptibility to a more preferred agent.

The model was structured to estimate the impact on mortality and LOS of reduction in the time to ID of the pathogen and of resistance mechanisms. Detection of pathogen with mRDT allows earlier optimisation of therapy. The model considers ID and subsequent actions on results at the species, group or genus level to be equivalent in the base case. In scenario analysis, the benefit of early ID is limited to species and group calls, or species calls only. The model considers three adverse outcomes arising from non-optimal treatment: increased mortality arising from a delay to effective treatment, risk of *C. difficile* infection, and risk of AKI arising from extended exposure to broad-spectrum antimicrobial therapy. The model considers the proportion of patients receiving effective empiric antimicrobial treatment according to the causative pathogen. Where empiric antimicrobial treatment is ineffective, patients are subject to a delay to effective treatment which increases the risk of mortality and extended hospital stay. After ID of the pathogen (and resistance pattern), patients receiving effective empiric antimicrobial treatment or unnecessary antimicrobial therapy may be de-escalated to a targeted therapy (or no therapy in the case of a contaminant). These patients are at heightened risk of *C. difficile* infection and AKI while on broad-spectrum antimicrobial therapy.

The model captures mortality, adverse events and costs over the index period of hospitalisation (assumed to be up to 30 days). Survivors of the index hospitalisation are assumed to recover completely to their previous health status (assumed to be age/sex matched to the general population). Quality adjusted life expectancy (QALE) of survivors is calculated assuming population data for mortality and health related quality of life (HRQoL). Patient HRQoL during hospitalisation is assumed to be unaffected by any reduction in time to ID of pathogens or resistance mechanisms. Where a patient has a pathogen with a resistance mechanism, changes to antimicrobial therapy are assumed to occur only after confirmation of the resistance mechanism. The key assumptions underpinning the analysis are described in Table 1.

**Table 1.**
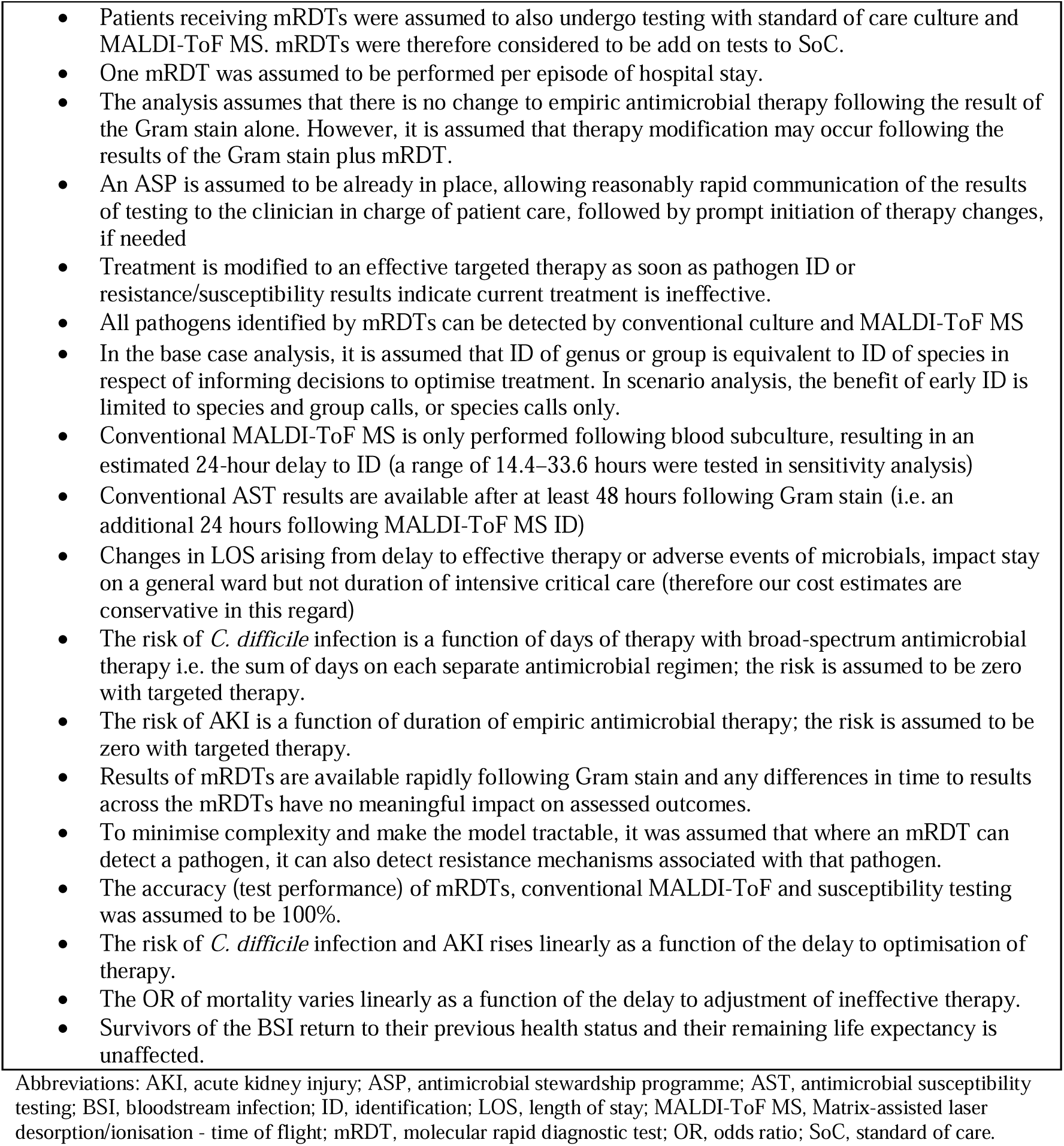
Main assumptions underpinning the analysis.

### 2.2 Intervention and comparators

The SoC for detection of BSIs is set to be conventional culture plus MALDI-ToF MS without mRDT. Conventional antimicrobial susceptibility testing was assumed to be undertaken using phenotypic AST methods such as disk diffusion, broth microdilution and automated AST systems such as VITEK^®^ 2, BD Phoenix^™^, or MicroScan^®^.

The interventions considered in the analysis are summarised in Table 2. All relevant panels (gram-positive, gram-negative and fungal) were included for each mRDT.

**Table 2.**
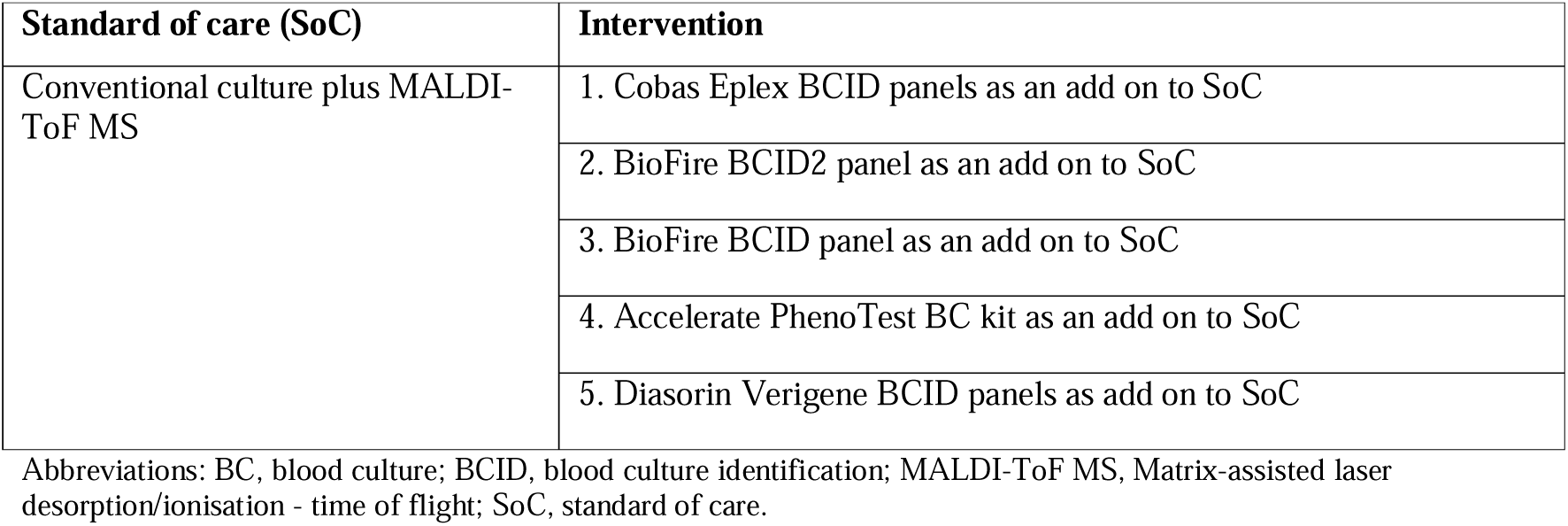
Standard of care and interventions.

### 2.3 Model parameters

A number of relevant parameters were sourced from a recent publication examining the relationship between receipt of appropriate antimicrobial therapy and mortality in 32,100 patients with a BSI across 183 US hospitals [22]. The study reported the proportion of patients for whom empiric antimicrobial therapy was effective across 17 broad species of pathogen. The effectiveness of empiric antimicrobial therapy ranged from 64.6% for *Acinetobacter baumannii* to 98.2% for *Staphylococcus aureus.* These data were used to determine the likelihood that empiric therapy is effective for each species of pathogen included in the analysis. The study also provided data on the relative proportion of gram-positive, gram–negative and fungal infections (46.59%, 52.50%, and 0.91%, respectively). An age of 65 years and a proportion of 45% women was assumed for the modelled cohort to align with the demographic data reported in the study. Finally, the study provided data on the number and class of antimicrobial therapies received by patients according to the type of BSI that was subsequently determined. Further details on the methods and data analysis are reported in the supplementary material.

The model considered 82 common BSI pathogens (27 Gram-negative, 37 Gram-positive, 15 fungal, and 3 generic categories for the remaining species within each pathogen type). Data on the distribution of pathogen species within the three types were derived from data collected during the development of the Cobas Eplex BCID panels. The pathogen species and genera detected by each mRDT were determined from the product information for each of the panels [23–27] (supplementary material, section S3.1.3). Pathogen species considered to be contaminants were guided by the literature [28] (supplementary material, section 3.2.2). The base case assumed no mortality associated with pathogens categorised as contaminants. However, in scenario analysis, all species were assumed invasive. The base case proportion of patients de-escalated from effective broad-spectrum antimicrobial therapy to a targeted therapy following ID of the pathogen or resistance mechanism was 62.9%, based on data at 72 hours for a tertiary care medical centre in the US [29].

Data on mortality were taken from a recent Canadian study that included 19,326 patients. The study reported 30-day mortality for 29 pathogen species [30]. Mortality ranged from 12.1% for *E. coli* to 41.9% for *Clostridium* species. These data were used to estimate mortality under SoC for each of the 82 pathogens in the model under the assumption that deaths from BSIs occur in the first 30-day period. The data are reported in detail in the supplementary material, section 3.1.6.

The impact of delays in receipt of effective therapy on mortality were taken from a recent Swedish study which reported odds ratios (OR) for mortality as a function of the delay to effective therapy based on analysis of 10,628 episodes of BSIs in adults [31]. A linear regression of the reported data estimated an increase in the OR of death of 0.01 for each hour of delay. Details of the analysis are provided in the supplementary material, section 3.1.7. The data were used to calculate the reduction in mortality associated with a reduction in time to ID of the pathogen or resistance mechanisms, where an mRDT was able to detect the pathogen and empiric therapy is ineffective.

The LOS associated with a BSI was taken from a US publication reporting LOS for 830 patients with a BSI according to whether empiric antimicrobial therapy was effective or not [32]. Digitisation of Kaplan-Meier (KM) data provided a restricted mean LOS of 11.56 days for patients receiving effective broad-spectrum empiric therapy. The impact of delay to effective therapy on LOS was parameterised using data from a US study which reported the impact of delay to Gram stain notification on LOS [33]. A duration ratio of 1.004 (95% confidence interval [CI]: 1.001, 1.008) was reported for each hour of delay. This ratio was used to increase LOS from the base value of 11.56 days for every hour of delay to ID of the pathogen or resistance mechanisms for patients whose empiric therapy is ineffective.

The risk of *C. difficile* infection was taken from a US study reporting the incidence of *C. difficile* infection as a function of the sum of defined daily doses of antibiotics received by the patient [34]. The defined daily dose is a dose considered sufficient for one day of treatment according to the World Health Organization (WHO), and hence the sum captures the cumulative impact of multiple antibiotic treatments. The study reported crude risks of *C. difficile* infection of 1.32% and 3.01% for defined daily doses in the range 3.0 to 7.79 and 7.8 to 21.0, respectively. Linear interpolation between the two data points generated an absolute risk of 0.19% of *C. difficile* for each defined daily dose of antibiotic. Mortality from *C. difficile* infections of 8.6% was taken from a study reporting mortality from *C. difficile* infection in patients with sepsis [35]. An increase in LOS of 3.66 days was assumed following a *C. difficile* infection [36]. The source was a systematic review which pooled estimates according to the rigour of the methodology in the respective studies; the chosen value was reported for studies using time-varying matching techniques.

The risk of AKI was taken from a US study in patients receiving vancomycin at doses of ≥4 g/day and <4 g/day, and patients receiving linezolid [37]. Data on time to nephrotoxicity for patients receiving < 4g/day of vancomycin indicated a cumulative risk of 16.7% at 12 days. Assuming that the risk of AKI is constant over time, this equates to a daily risk of 1.51%. The model assumed a reduction in risk of AKI of 1.51% for every 24-hour reduction in the duration of broad-spectrum antimicrobial therapy. The impact of AKI on mortality was taken from a recent US study examining the incidence and outcome of AKI in patients with carbapenem-resistant Gram-negative infections [38]. Adjusted mortality was 18.5% in the group who experienced AKI (n=91) compared with 5.6% in the group who did not (n=659). The absolute difference of 12.9% was assumed as the mortality attributable to AKI. The impact of AKI on LOS was taken from a US study examining the incidence and severity of AKI associated with antimicrobial therapy [39]. The study reported median and interquartile range (IQR) for LOS in patients not experiencing and experiencing AKI of 11 (IQR: 8, 17) and 15 (IQR: 11, 23), respectively. The median LOS were converted to mean LOS of 11.53 and 15.84 days using the method of Wan et al. [40]. The difference between the mean values of 4.31 days was assumed to be the increase in LOS associated with AKI.

Data on the classes of empiric therapy received by patients (varying by class of pathogen) were taken from Ohnuma et al [22]. Expert clinical opinion was sought to identify the most likely antimicrobial regimen used for each of the classes of antimicrobial, and for the most likely targeted therapy for each pathogen species in the model. Data on dosing were primarily obtained from the British National Formulary [41] and combined with unit costs from the US Veteran Affairs drug pricing database [42] to allow estimation of the costs of broad-spectrum and targeted antimicrobial therapies (supplementary material, section 3.2.2). The data allowed estimation of the cost impact of moving from broad-spectrum to targeted therapy.

The cost of mRDTs were estimated using Roche market analysis. The costs include reagents, rental and maintenance of the machine for a typical mid-sized hospital. The cost of MALDI-ToF MS did not impact the analysis since mRDTs were assumed to be used in addition to SoC (culture and MALDI-ToF MS). A cost of $7.52 per sample, which included reagents and capital costs, was estimated based on data reported by Tran et al. [43] (supplementary material, section 3.2.1). A cost of hospital stay per inpatient day of $3,025 was taken from data by the Kaiser Family Foundation (KFF) [44]. This is an all-inclusive cost of inpatient expenses including the cost of antimicrobials. Costs are reported in 2024 US Dollars (USD).

Key model parameters are summarised in Table 3.

**Table 3.** Key model parameters.

| Parameter | Value(s) | Source |
| --- | --- | --- |
| Patient demographics | Age 65, 45% female | Ohnuma 2023 [22] |
| Distribution of pathogens by class (US) | Gram-negative – 46.6%; gram-positive 52.5%; fungal 0.9% | Ohnuma 2023 [22] |
| Distribution of pathogens by class (UK) | Gram-negative – 41.7%; gram-positive 57.3%; fungal 1.0% | Public Health England [45] |
| Effectiveness of empiric therapy | 64.6% to 98.2% according to pathogen species | Ohnuma 2023 [22] |
| Thirty-day mortality | 12.1% to 41.9% according to pathogen species | Verway 2023 [30] |
| Impact of delay to effective therapy on mortality | OR increased by 0.01 for each hour delay | Van Heuverswyn 2023 [31] |
| LOS for patients receiving effective empiric therapy | 11.56 days | Battle 2016 [32] |
| Impact of delay to effective therapy on LOS | Duration ratio of 1.004 for each hour delay | Beekman 2003 [33] |
| Risk of *C. difficile* infection | 0.19% per defined daily dose of antimicrobial | Stevens 2011 [34] |
| Mortality associated with *C. difficile* infection | 8.6% | Lagu [35] |
| LOS associated with *C. difficile* infection | 3.66 days | Manoukian 2018 [36] |
| Risk of AKI | 1.51% per day in receipt of broad-spectrum antimicrobial therapy | Lodise 2008 [37] |
| Mortality associated with AKI | 12.9% | Lodise 2024[38] |
| LOS associated with AKI | 4.31 days | Minejima 2011 [39] |
| Delay to pathogen ID with MALDI-ToF compared with mRDT | 24 hours | Assumption |
| Delay in receiving AST /resistance results compared to mRDT | 48 hours | Assumption |
| De-escalation from effective broad-spectrum therapy following pathogen ID | 62.9% | Liu 2016 [29] |
| Cost of mRDT | \$64 to \$115 | Roche market analysis |
| Cost of antimicrobials (per day) | \$3.11 to \$1,236.38 | Veteran affairs drug pricing database [42] |
| Cost of bed day in hospital (US) | \$3,025 | Kaiser Family Foundation [44] |
| Cost of bed day in hospital (UK) | £933 | Manoukian 2018 [36] |

Abbreviations: AKI, acute kidney injury; AST, antimicrobial susceptibility testing; ID, identification; LOS, length of stay; MALDI-ToF, Matrix-assisted laser desorption/ionisation - time of flight; mRDT, molecular rapid diagnostic test; OR, odds ratio; UK, United Kingdom; US, United States.

Life years accrued for survivors were estimated using US national life tables for 2020 [46]. Life years were discounted at 3%. Life years were weighted for HRQoL using utility data as a function of age and sex for the US population to estimate QALYs [47].

### 2.4 Sensitivity analysis

Probabilistic sensitivity analysis (PSA) was undertaken to quantify the impact of uncertainty across model parameters that were subject to sampling uncertainty. Distributions were assigned to each parameter reflecting natural bounds on plausible parameter values with standard errors estimated from the original parameter source, where possible, or assumed to be ±20% of the mean value. One thousand simulations of the model were run with each parameter value sampled from the specified distribution in each simulation. Results are reported as the cost-effectiveness acceptability curve.

One-way sensitivity analysis (OWSA) was performed on all parameters subject to sampling uncertainty (with the exception of life-table data and data on HRQoL as a function of age), with CIs informed by standard errors, where these data were available, or estimated as ±20% of the point estimate.

Scenario analysis was conducted to examine cost-effectiveness in a UK setting. In this scenario, the distribution of pathogens, LOS in hospital, and unit costs were estimated from sources relevant to the UK. Scenario analyses were also undertaken in which genus calls, or both genus and group calls were excluded (treated as not identifying the pathogen). Additionally, scenario analysis was undertaken in which all pathogens were assumed to be true infections rather than contaminants. Scenario analysis was also undertaken to examine the impact of assuming a lower proportion of patients are de-escalated from effective antimicrobial therapy to targeted therapy following pathogen or resistance ID, given the heterogeneity of practice with regard to de-escalation [48].

## 3 Results

### 3.1 Base case analysis

Table 4 presents the base case results. All mRDTs, as add on to SoC, generated lower costs than conventional culture and MALDI-ToF MS (SoC) alone. The lowest costs were associated with the Cobas Eplex BCID panels, which generated cost savings of $164 compared with the SoC. All strategies incorporating a mRDT generated improved outcomes compared to the SoC. Cobas Eplex BCID panels generated the highest QALYs and the fewest deaths across comparators, saving 24 lives per 10,000 patients and generating 0.024 additional QALYs compared with SoC. Consequently, Cobas Eplex BCID panels dominated all other comparators (i.e. they were more effective and generated lower costs).

**Table 4.** Summary of costs, QALYs, and mortality.

| Diagnostic strategy | Overall cost (per patient) | QALYs (per patient) | Deaths (per 10,000 patients) |
| --- | --- | --- | --- |
| Conventional culture and MALDI ToF MS (SoC) | \$35,515 | 8.917 | 1127.8 |
| Diasorin Verigene BCID panels | \$35,365 | 8.934 | 1111.1 |
| Accelerate PhenoTest BC kit | \$35,456 | 8.937 | 1108.3 |
| BioFire BCID panel | \$35,372 | 8.939 | 1105.9 |
| BioFire BCID2 panel | \$35,364 | 8.940 | 1104.7 |
| Cobas Eplex BCID panels | \$35,351 | 8.941 | 1103.6 |

Abbreviations: MALDI-ToF MS, Matrix-assisted laser desorption/ionisation - time of flight mass spectrometry; pop, population; QALY, quality-adjusted life year; SoC, standard-of-care.

Figure 1 presents a breakdown of the mortality gains for each mRDT compared with SoC. For the majority of mRDTs, reduction in time to effective therapy for patients receiving ineffective empiric therapy was responsible for more than half of the total reduction in mortality. The exception was Diasorin Verigene BCID panels, where mortality gains from AKI averted exceeded gains from reduction in time on ineffective empiric therapy. The latter result reflects high mortality and lower effectiveness of empiric therapy for fungal pathogens combined with the lack of any fungal organisms being detected on the Diasorin Verigene BCID panels. Reduction in the incidence of *C. difficile* infection was the smallest contributor to mortality gains.

**Figure 1.**
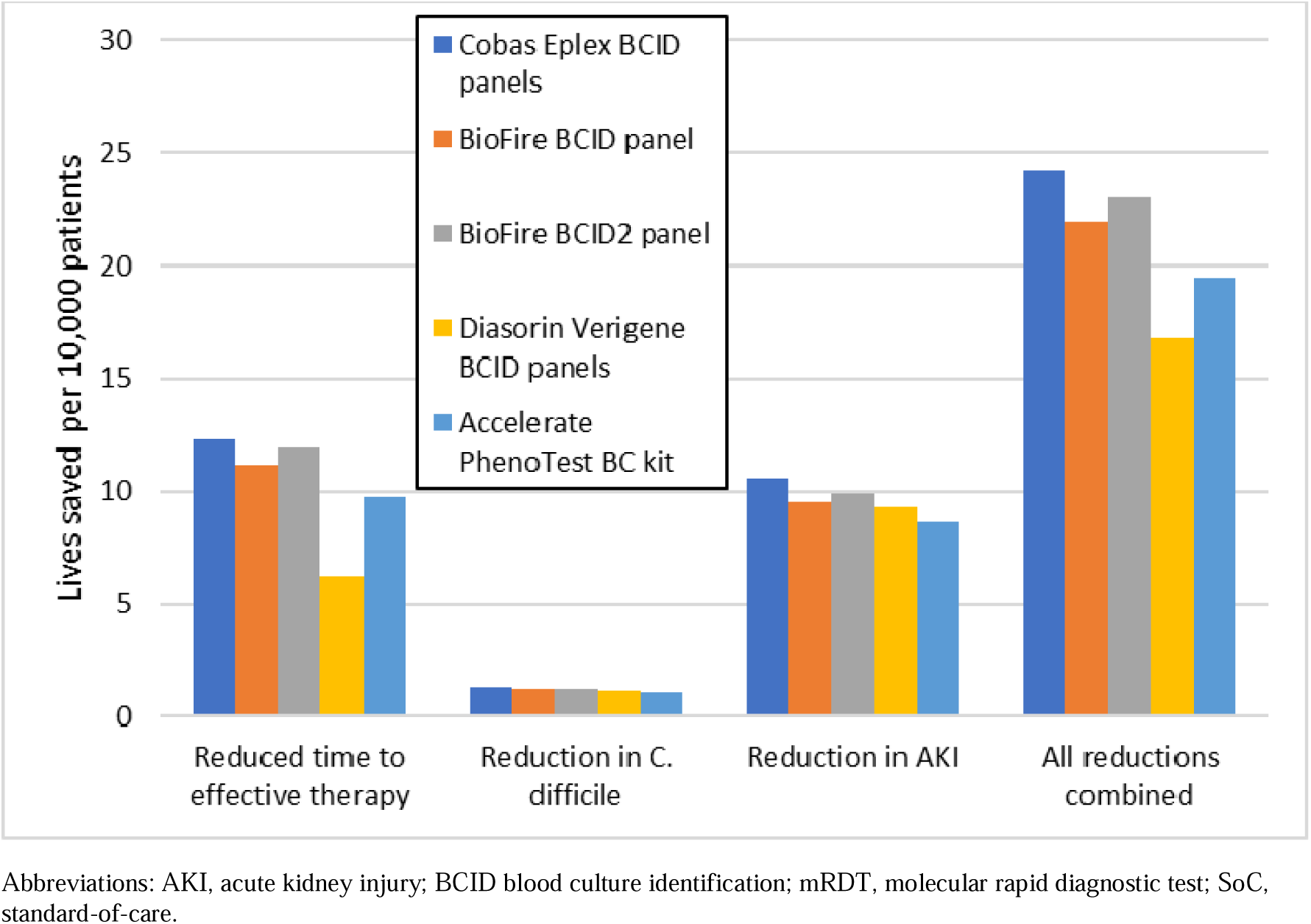
Breakdown of deaths averted by category for mRDT compared with SoC

A breakdown of the cost savings associated with mRDTs strategies compared with SoC is shown in Figure 2. Analogous to the reductions in mortality, the largest contributor to the cost savings for most mRDTs is the reduction in LOS from reduced time to effective therapy in patients receiving ineffective empiric therapy. The next largest cost saving arises from reduction in LOS from reductions in AKI. There is a modest saving in antimicrobial costs arising from earlier switching from broad-spectrum to targeted therapy.

**Figure 2.**
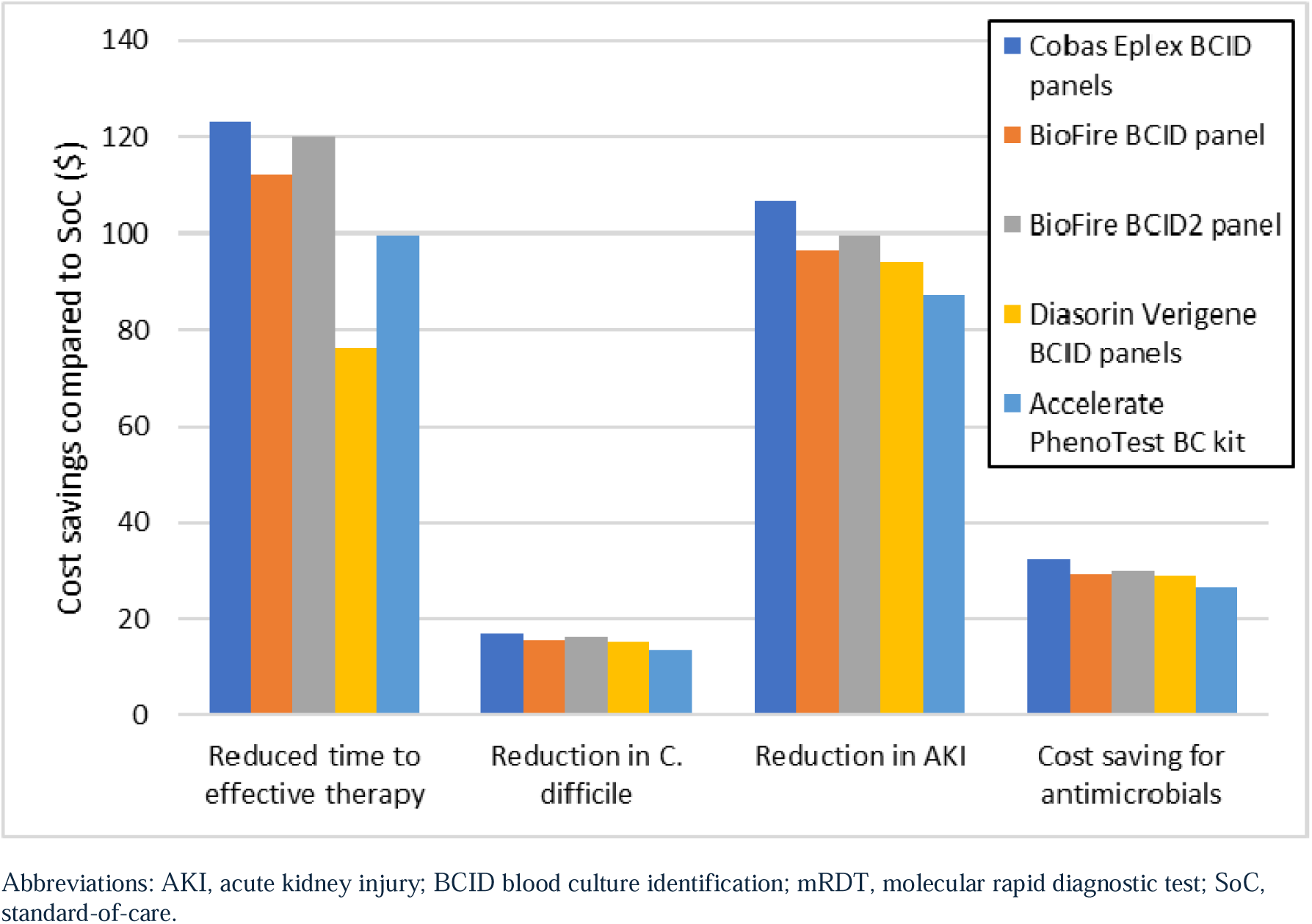
Breakdown of cost savings (per patient) for mRDTs strategies compared with SoC

### 3.2 Sensitivity and scenario analyses

The cost-effectiveness acceptability curves (CEAC) for Cobas Eplex BCID panels, Diason Verigene BCID panels and BioFire BCID2 with respect to QALYs are presented in Figure 3. Curves for SoC, Accelerate PhenoTest BC kit and BioFire BCID1 are <1% across the range of willingness-to-pay values for a QALY indicating that they are never cost-effective. (The curves are omitted from Figure 3 for clarity.) The probability that the Cobas Eplex BCID panels are cost-effective exceeds 98% at a value of $10,000 per QALY.

**Figure 3.**
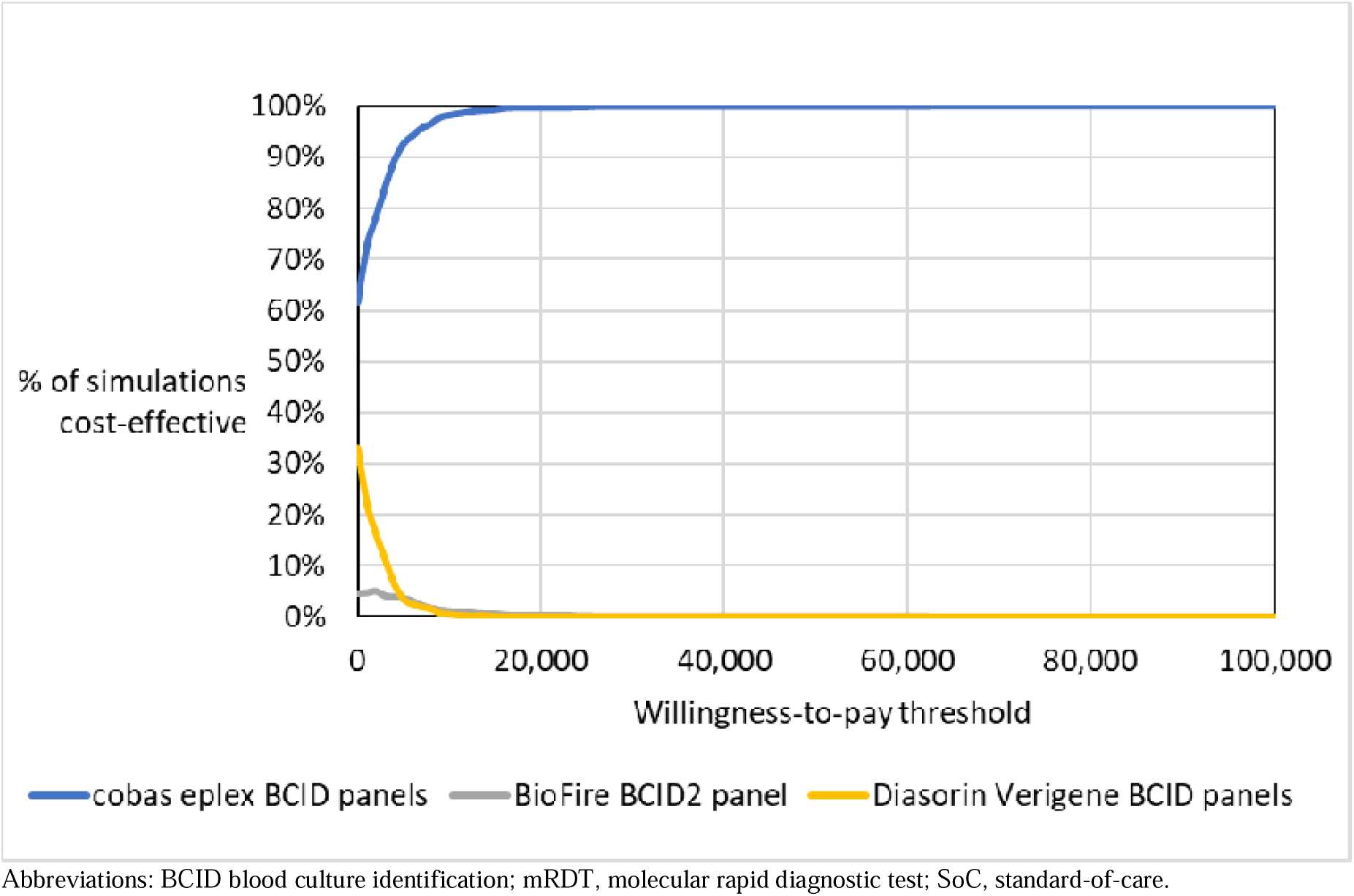
Cost-effectiveness acceptability curve for Cobas Eplex BCID panels, Diasorin Verigene BCID panels and BioFire BCID2 panel

Figure 4 presents the OWSA for Cobas Eplex BCID panels compared with BioFire BCID2 panel, the next most effective mRDT. Results are presented in terms of the net monetary benefit (NMB) to avoid challenges in the interpretation of negative incremental cost-effectiveness ratio (ICERs) that arise when a technology is dominant. A conservative value of $50,000 per QALY was applied. The parameter with the largest impact on the cost-effectiveness of the Cobas Eplex BCID panels compared with BioFire BCID2 panel is the time to ID of pathogens with SoC. This parameter influences the costs and benefits of early detection for each pathogen detected by either mRDT. The second most influential parameter is the age of the cohort. This parameter influences the benefits of early detection of any pathogen. The cost of a test with Cobas Eplex BCID panels and BioFire BCID2 panel are the third and fourth most influential parameters. Cobas Eplex BCID panels remain cost-effective in all OWSA. Additional OWSA is presented in the supplementary materials, section 4.0.

**Figure 4.**
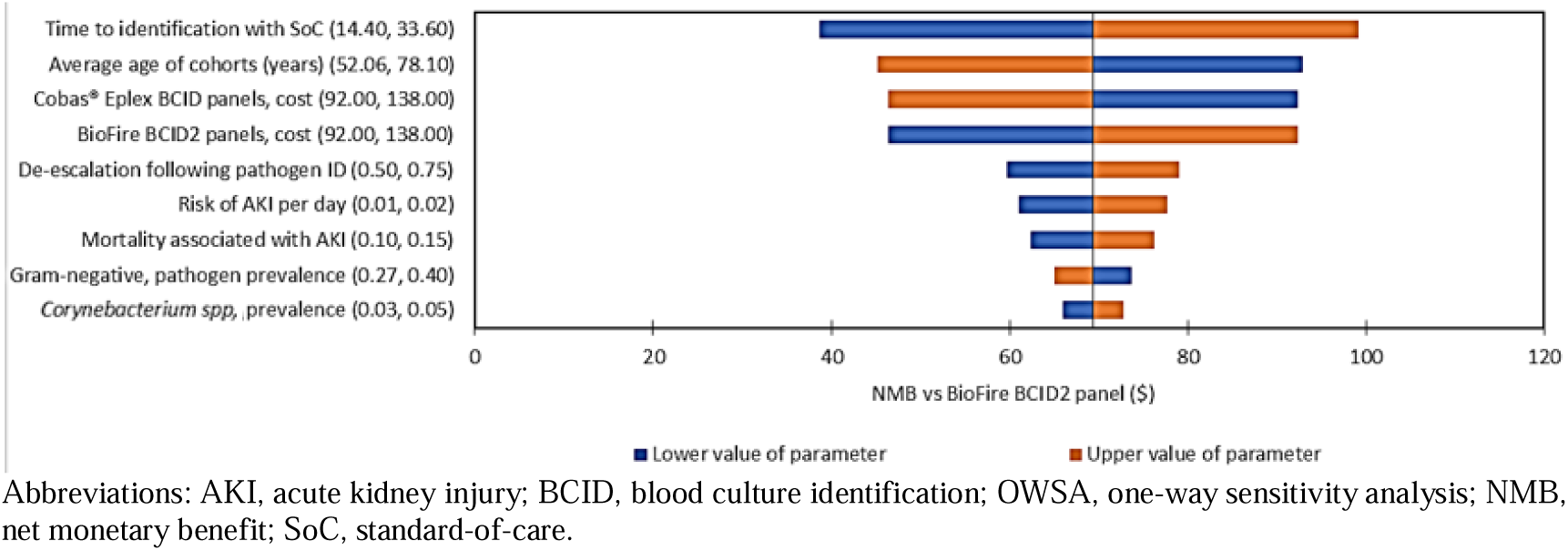
Tornado plot for the OWSA of Cobas Eplex BCID panels versus BioFire BCID2

In the scenario analysis considering a UK setting, the cheapest comparator was Diasorin Verigene BCID panels. Cobas Eplex BCID panels was the most effective comparator with an ICER of £14.190 per QALY. In all other scenario analyses in a US setting, Cobas Eplex BCID panels remained dominant over all other comparators (details in the supplementary analysis). At a willingness-to-pay of $50,000 per QALY, Cobas Eplex BCID panels remained cost-effective down to a test cost of $46 for BioFire BCID2 panel. Scenario results are reported in detail in the supplementary materials, section 4.1.

## 4 Discussion

### 4.1 Key findings

This study evaluated the cost-effectiveness of adding mRDTs to the established SoC method for diagnosing BSI (i.e. conventional culture plus MALDI-ToF MS). The primary comparator was SoC alone with no mRDTs, and overall, all mRDT strategies were cost saving. This result is unsurprising in that earlier detection of pathogens and antimicrobial resistance genes, and more rapid optimisation of effective antimicrobial therapy, would be expected to improve outcomes in patients whose empiric antimicrobial therapy is ineffective, and this in turn would be expected to reduce LOS. Given the high cost of a single bed day in a US hospital, only a very modest reduction in LOS (of about an hour) is required to offset the cost of procuring the mRDTs. The analysis also demonstrates that Cobas Eplex BCID panels are cost-effective when compared with other mRDTs. The additional panel coverage of the Cobas Eplex BCID panels compared with other mRDTs is sufficient to offset any additional test costs and ensure that the Cobas Eplex BCID panels is cost-effective. The Cobas Eplex BCID panels remains cost-effective even if the price of the next most effective mRDT, BioFire BCID2 panel, is reduced to $50 in the US setting.

Alongside escalation of therapy in cases of ineffective (e.g. narrow) empiric antimicrobial therapy, de-escalation from broad-spectrum antimicrobial therapy is important to prevent adverse events and limit the spread of antimicrobial resistance. This analysis considered the additional time required for ID and AST which can further delay time to effective therapy with conventional MALDI-ToF MS compared with mRDTs. It also considered two of the most serious adverse events from broad-spectrum antimicrobial treatment, *C. difficile* infection and AKI. Cost savings from reductions in AKI and *C. difficile* infection, in addition to cost savings from de-escalation of antimicrobials were more than sufficient to offset the cost of procuring the mRDTs. These results highlight the economic benefits of de-escalation, in addition to the benefits with respect to antibiotic stewardship.

The burden of mortality from BSIs remains considerable [30]. The increasing threat posed by resistant pathogens raises the possibility that the burden may increase over time. The advantage of mRDTs in combating this is two-fold. Firstly, more rapid ID of pathogens reduces mortality. Second, and possibly more importantly, when combined with an effective antibiotic stewardship program and de-escalation of broad-spectrum therapy, mRDTs can help to slow the spread of resistant pathogens. Wide coverage of pathogens is important to maximise the effectiveness of mRDTs. The Cobas Eplex BCID panels provide the broadest coverage of BSI organisms and their resistance markers, including anaerobes and multi-drug-resistant organisms, pathogens which can be considered contaminants, along with common and emerging fungal pathogens. This comprehensive coverage generated lower costs alongside improved patient outcomes when compared to other mRDTs as well as SoC.

A number of previous economic evaluations have evaluated mRDTs compared with conventional methods and concluded that mRDTs are dominant [16, 17, 49–52]. However, previous evaluations are subject to a number of limitations including a lack of differentiation of mortality for BSI across different pathogens or differentiation of coverage across different mRDTs. In particular, previous evaluations have applied arbitrary assumption of higher mortality and higher LOS with the use of conventional methods [17], or with ineffective empiric therapy [50–52].

In contrast, the current analysis sought to improve on existing methods in the following ways: the model considers a population with suspected BSI; captures the impact of empiric antimicrobial therapy; and considers the reduction in time to effective therapy associated with mRDTs, by pathogen type, and relates this to changes in mortality (by pathogen type) and LOS. Therefore, the current model allows comparison between mRDTs on the basis of coverage, alongside a comparison with MALDI-ToF MS, and allows flexibility to consider the impact of changes in the distribution of microbiota on the effectiveness and cost-effectiveness of different mRDTs.

### 4.2 Strengths and limitations

This analysis has a number of strengths. The analysis considers the evidence on the current care pathway, notably, the proportion of patients receiving effective empiric therapy. This aspect is critical as the vast majority of patients receive effective empiric therapy. The analysis utilised recently published data which would have been unavailable to authors of previously published economic analyses. These data allowed the analysis to capture the much higher probability that empiric therapy will be ineffective in some less common pathogens, and to reflect differences in mortality across pathogen species. The model directly links time to effective therapy to improvements in LOS and mortality, the latter utilising a recent publication which showed a clear and broadly linear relationship between time to effective therapy and the OR of death [31]. The model considered pathogen resistance and the delay to effective therapy resulting from the need for susceptibility testing in the conventional pathway. The model allows comparison of different mRDTs according to the pathogens they detect, reflecting the prevalence and severity of those pathogens.

Some simplifications were imposed on the analysis to keep the model tractable. The model did not consider the detection of resistance mechanisms separately to the detection of pathogen species. Instead, a simplifying assumption was made that a mRDT capable of detecting a pathogen could also detect common resistance mechanisms. The model considered only two adverse events of broad-spectrum antimicrobials: *C. difficile* infection and AKI. The model did not consider the potential savings arising from reduced need for critical care as a result of earlier pathogen detection. In this respect, the results of the model can be considered conservative. The model was subject to some limitations in the available data to support the analysis. Evidence on the time taken for conventional culture and MALDI-ToF MS, and the time savings associated with mRDTs was disparate. Conventional MALDI-ToF MS requires pure bacterial colonies, and an assumption was made that the sub-culturing process and growth takes 24 hours. Sensitivity analysis in which a time of 14.4 hours was assumed in place of 24 hours generated the lowest NMB for the Cobas Eplex BCID panels compared with the SoC, but the Cobas Eplex BCID panels remained cost-effective (details in supplementary material, section 4.1). The impact of time to effective therapy on LOS was taken from an old study which reported the impact of the time to Gram stain results on LOS.

It is notable that the largest trial comparing MALDI-ToF MS with conventional culture failed to demonstrate the cost-effectiveness of MALDI-ToF MS [53]. Whilst the cost-effectiveness of the Cobas Eplex BCID panels compared to MALDI-ToF MS has been demonstrated with respect to median time to optimised therapy [54], it would be challenging to demonstrate the cost-utility of the Cobas Eplex BCID panels with MALDI-ToF MS or another mRDT in a clinical trial due to the modest differences in mortality across comparators. These differences are important (particularly to the relevant survivors), but differences of the magnitude of 0.5% to 0.05% are challenging or infeasible to measure with confidence in a trial. Yet the differences are sufficient to justify the cost of mRDTs. This analysis demonstrates that in general, mRDTs are cost-effective compared with MALDI-ToF, and that the Cobas Eplex BCID panels is cost-effective compared with other mRDTs.

### 4.3 Conclusion

This analysis evaluated the cost-effectiveness of commonly available mRDTs as add-on tests to the standard culture and MALDI-ToF MS techniques. The analysis considered the coverage of each mRDT and the impact of reduced time to pathogen ID on mortality and LOS. The analysis found that any approach that included mRDT dominated SoC, and given the broader pathogen coverage, the Cobas Eplex BCID panels dominated other mRDTs. The findings of the analysis are robust to both OWSA and PSA. However, they are dependent on assumptions regarding the rapid availability of mRDTs results following a Gram stain, conveyance of information to the treating clinician, and the assumption the clinician would act rapidly to optimise therapy.

## Supporting information

CHEERS statement

## Data Availability

The analysis used secondary data sources which are publicly available. No primary individual patient data was collected or used.

## Supplementary material

## Notes

### Author Declarations

The analysis used secondary data sources. No individual patient data was collected or used. Consequently, ethical approval for the study was not required

