## Supplementary material for "Evaluating the cost-effectiveness of rapid diagnostic testing for the identification of pathogens and resistance genes in bloodstream infections"

TITLE

RUNNING TITLE

Cost-effectiveness of rapid diagnostic tests for suspected blood-stream infection

James K. Karichu^1^, Mark Pennington^2^, Xiyu Bao^2^, Kiera Lander^2^, Tiffeny T. Smith^3^, Adam Thornberg^1^

1. Roche Molecular Systems, Pleasanton, California, USA
2. Source Health Economics, London, UK
3. Roche Diagnostics Corporation, Indianapolis, Indiana, USA

Name and active e-mail address of the corresponding author

Name: James K. Karichu

Department/Institution: Global Access & Policy, Roche Diagnostic Solutions

Address:4300 Hacienda Drive, Pleasanton, CA, 94588

James K. Karichu: 0000-0002-2052-4659

Mark Pennington: 0000-0002-1392-8700

Tiffeny T. Smith: 0009-0005-2426-4427

Adam Thornberg: 0009-0004-8359-6336

Supplementary material

1. Clinical pathway and position of molecular rapid diagnostic tests
   1.1. Model conceptualisation

A formal health economic analysis plan was not developed. However, model development followed a conceptualisation phase in which input was sought from clinical experts involved in the project. The starting point was the assumption that rapid molecular diagnostic tests (mRDT) provide earlier identification of the causative pathogen of a blood stream infection (BSI) compared with conventional matrix-assisted laser desorption ionization–time of flight mass spectrometry (MALDI-ToF MS). From this, stems the axiom that mRDT offer no benefits to patients with a BSI that is not detected by the technology. A further assumption was made that, for at least some patients, clinicians act on the results of the diagnosis to change how the patient is managed. This assumes relatively rapid implementation of the mRDT and communication of the results.

A simplification of the care pathway is shown in Figure 1. The circles show the key steps in the initial treatment of BSI up to the point of definitive identification of the causative pathogen. Rectangles indicate changes in the care of the patient arising from diagnostic information. Rounded squares represent the current treatment status of the patient. Hexagons represent the impact of diagnostic information on costs and outcomes for patients. The pathway considers a patient admitted to hospital with a suspected BSI. The patient is assumed to be rapidly started on intravenous (IV) broad-spectrum antimicrobial therapy. This decision, and the composition of the therapy will be informed by the patient case history, presentation, the local hospital treatment guidelines (particularly with regard to selection of antimicrobials), and the judgement of the treating clinician. Prior to commencement of empiric therapy, blood samples are drawn for culture to facilitate identification of the causative pathogen.

Figure 1: Conceptualisation of the care pathway for the diagnosis and initial treatment of BSI

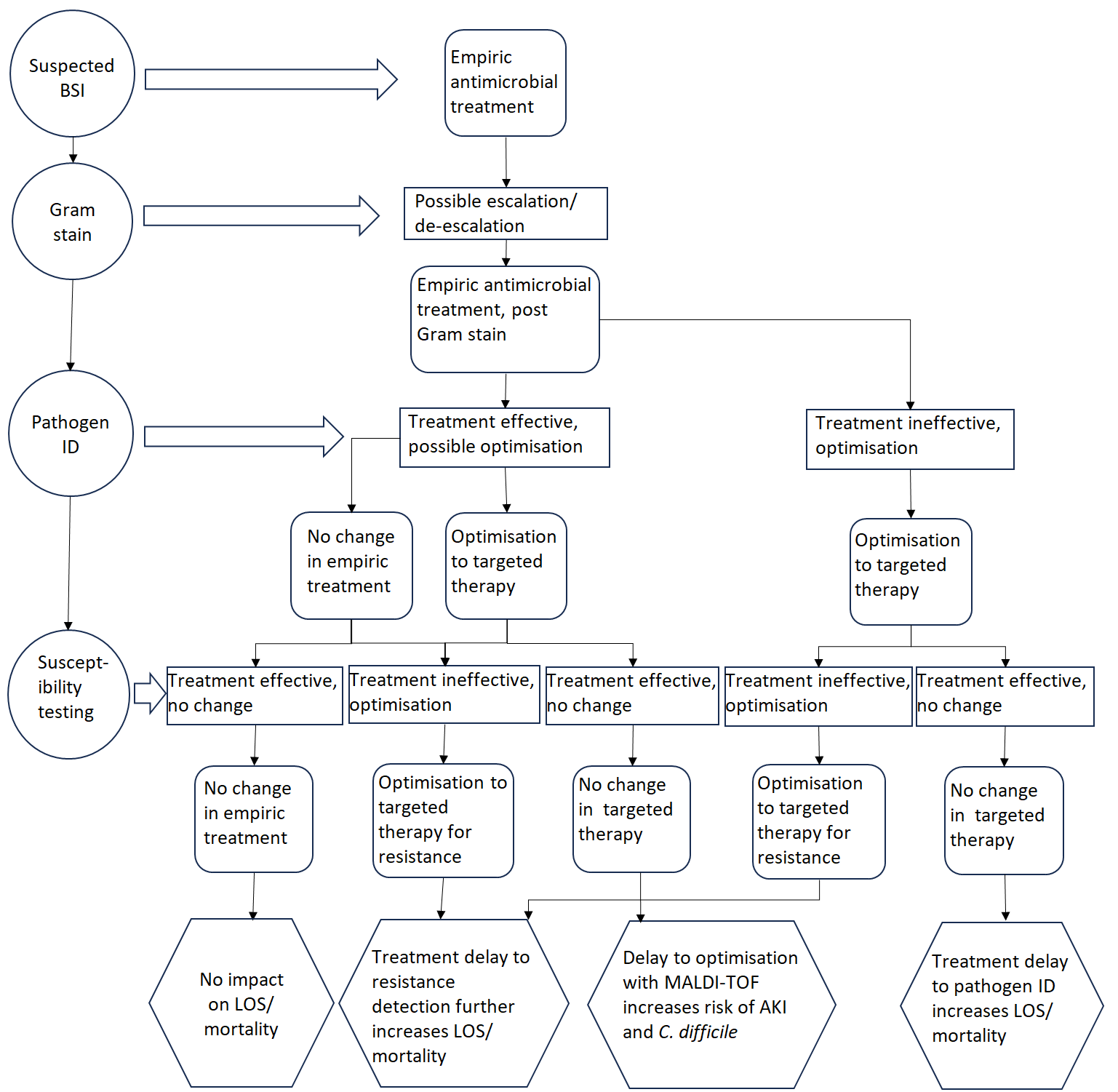

Circles show the key steps in the initial treatment of BSI up to the point of definitive identification of the causative pathogen. Rectangles indicate changes in the care of the patient arising from diagnostic information. Rounded squares represent the current treatment status of the patient. Hexagons represent the impact of diagnostic information on costs and outcomes for patients.
Abbreviations: AKI, acute kidney injury; BSI, bloodstream infection; ID, identification; LOS, length of stay; MALDI-ToF, matrix-assisted laser desorption/ionisation - time of flight mass spectrometry.

The first step in the blood culture process is to draw two sets of blood culture bottles (aerobic and anaerobic) from two separate venipuncture sites, and these bottles are incubated in continuous monitoring systems for at least 5 days or until they ring positive, whichever comes earlier. Once a bottle records as positive, a Gram stain is set up immediately (plus a mRDT, if available). Gram stain provides information on the morphology of the pathogen: gram-positive bacteria, gram-negative bacteria, or fungal pathogen. Clinicians may act on this information to optimise therapy. Following the Gram stain, the process of identification of the pathogen is commenced. The conventional approach requires further culturation of the blood sample (sub-culture on solid agar plates followed by 18–24-hour incubation) to provide pure colonies to allow identification by MALDI-ToF MS and antimicrobial susceptibility testing (AST).

As noted above, if available, a rapid mRDT may be undertaken to allow faster identification of the pathogen, and resistance mechanism(s). Identification of the pathogen (plus resistance profile) allows the treating clinician to determine whether empiric therapy is effective against the type of pathogen or whether a change is needed. Clinicians would be expected to change therapy to an effective antimicrobial (escalate) if the current empiric therapy is ineffective against the pathogen. If the current empiric therapy is effective against the pathogen, the clinician may elect to change to a targeted therapy (optimise). Optimisation typically happens if the detected pathogen is susceptible to narrower-spectrum options. For instance, if *Escherichia coli* with no resistance genes is detected, the clinician can optimise therapy from broad-spectrum piperacillin-tazobactam and vancomycin to a more targeted agent like ceftriaxone, which is effective against *E. coli* and has a narrower spectrum. This adjustment reduces the risk of adverse effects and unnecessary broad-spectrum antibiotic use.

Following identification of the pathogen using conventional MALDI-ToF MS, a further step is required to test the susceptibility of the pathogen to antimicrobials (AST). AST is reported in terms of the minimum inhibitory concentration (MIC) required to prevent growth of the pathogen. These results indicate which antimicrobials are likely to be effective against the pathogen and allow a decision either to escalate to an effective therapy from an ineffective therapy, or to de-escalate from an effective broad-spectrum therapy to a targeted therapy. This process requires additional time following pathogen identification for conventional MALDI-ToF MS. Whilst mRDT do not provide data on MIC, they can identify common resistance mechanisms which would render certain antimicrobials ineffective. This information is available at the same time as data identifying the pathogen. To simplify the modelling process, identification of a resistance mechanism for a specific antibiotic and data on MIC for that antibiotic are assumed to provide the same level of guidance on the appropriateness of the therapy.

Delays in providing effective therapy for BSIs are associated with poorer outcomes for patients [1]. Delays are also associated with increased length of stay (LOS) in intensive care units and overall hospital stay [2]. Hence more rapid identification of the pathogen and resistance mechanisms allows a more rapid escalation of therapy in patients whose empiric therapy is ineffective, with potential gains to patients in reducing mortality and to institutions in reducing LOS. Broad-spectrum antimicrobial therapy is also associated with adverse events [3]. The two adverse events considered most significant were *Clostridioides difficile* infection and acute kidney injury (AKI). Risks of both adverse events (AE) were considered to be mitigated by optimisation from broad-spectrum to targeted antimicrobial therapy. More rapid identification of pathogens and resistance mechanisms allows earlier optimisation to targeted therapy with the potential to reduce the risk of *C. difficile* infection or AKI.

Escalation or optimisation of therapy following identification of the pathogen or resistance mechanism depends on rapid communication of the results to the clinician, and potentially on clinical direction regarding appropriate changes to therapy. Antimicrobial stewardship programmes (ASP) are intended to provide guidance to clinicians on escalation or optimisation of therapy with the aim of reducing adverse outcomes for patients and the development of antibiotic resistance [4]. Programme composition varies, but typically includes a multidisciplinary team consisting of an infectious disease physician and a clinical pharmacist, along with infrastructure for the rapid communication of test results. There is evidence to indicate that early identification of pathogens has little impact on patient care in the absence of an ASP [5].

2. Model methods

The cost-effectiveness model (CEM) was designed to capture the incremental benefit of each pathogen detected by mRDT. The model captured the impact on patient mortality and LOS of the following changes in care pathway:

- Reduction in time to effective therapy (for patients whose empiric therapy is ineffective). Effective therapy is achieved by timely adjustment of antimicrobial treatment—either by broadening, or narrowing empiric therapy—based on diagnostic and clinical findings to ensure that the therapy targets the causative pathogen, minimises unnecessary antibiotic use and optimises patient outcomes. See examples in the Appendices.
- Reduction of *C. difficile* infection through early optimisation (de-escalation) to targeted therapy
- Reduction of AKI through early optimisation (de-escalation) to targeted therapy.

Patients entering the model fall into one of two groups with regard to empiric therapy: those for whom empiric therapy is effective against their BSI; and those for whom it is not. The model assumes that clinicians are rapidly informed of the result of the pathogen identification (for instance, an ASP is already in place). The model assumes that clinicians act rapidly to adjust therapy to an effective targeted treatment in patients receiving ineffective empiric therapy. The model assumes that clinicians act rapidly to optimise empiric therapy to a targeted therapy *for a proportion* of patients in whom empiric therapy is effective. These patients benefit from a reduction in the risk of AEs associated with the move from broad-spectrum empiric therapy to targeted therapy. The model evaluates the impact on mortality and LOS of the reduction in time to effective therapy. The model includes the cost of antimicrobials. The model evaluates the reduction in incidence of *C. difficile* infection and AKI as a function of the reduction in time on broad-spectrum antimicrobials in patients optimised from empiric therapy (in those patients whose empiric therapy is effective). The model considers bacterial species that are contaminants; patients are assumed to be optimised to no antimicrobial therapy instead of a targeted antimicrobial agent where a contaminant is identified. All patients are assumed to undergo testing using conventional culture plus MALDI-ToF MS in addition to any mRDT, if applicable. The treatment decisions for patients with a BSI that is not detected by an mRDT are assumed to be unaffected.

Patients with a BSI caused by a pathogen with a resistance mechanism are modelled in the same way as for patients with a BSI caused by a non-resistant pathogen. However, the time saving through rapid detection of the resistance mechanism by the mRDT compared to conventional culture and MALDI-ToF MS is larger as the latter requires the results of phenotypic AST to identify which antimicrobials are effective against the pathogen.

- In the absence of mRDT results and in patients with non-resistant pathogens, any changes to care are assumed to occur after receipt of MALDI-ToF MS pathogen identification.
- In the absence of mRDT results and in patients with a resistant pathogen, any therapy optimisation changes are assumed to occur after receipt of AST results; no change is assumed to occur following MALDI-ToF MS pathogen identification. Of note, currently MALDI-ToF MS is primarily used routinely for rapid organism identification, not for directly determining antibiotic resistance profiles. However, this capability is being explored.
- In reality, the pathway of care for patients receiving conventional culture, MALDI-ToF MS and susceptibility testing cannot be different for patients with and without a resistant pathogen prior to the point at which AST results are available. In effect, in the model, any changes to therapy following MALDI-ToF MS are ignored in patients who have a resistant infection. AST results, that can inform therapy optimisation decisions, are assumed to be available after 48 hours, in the base case analysis.

The model makes a number of further assumptions to reduce the complexity of modelling the care pathway. The impact on AEs of more rapid escalation from ineffective broad-spectrum therapy to effective therapy with broader coverage is ignored. In practice, patients may benefit from a reduction in risk of adverse events, alongside the benefit of effective antimicrobial treatment, when escalated from ineffective empiric therapy. An example here is when a patient is placed on ceftriaxone or piperacillin-tazobactam empiric therapy but an extended-spectrum beta-lactamase (ESBL)-producing Enterobacteriaceae is identified. Escalation to carbapenems may be needed because these antibiotics may not cover ESBL-producing *Escherichia coli* or *Klebsiella*. Modelling each permutation of pathogen and resistance mechanism would have been prohibitively complex. Consequently, the model assumed that where a mRDT detected a pathogen, it also detected resistance mechanisms relevant to that pathogen.

The model ignored any impact on short-term (30 days) quality of life of changes in the care pathway, only the impact on 30-‍day mortality was captured. Whilst the impact of BSI on patient quality of life during treatment is likely to be substantial, the duration of the impact renders the overall QALY loss small. Any long-term impact of a BSI on quality of life was ignored in order to maintain the simplicity of the model. The model also assumed that patients who survive 30 days make a full recovery from their BSI. Finally, the model assumed complete (100%) sensitivity and specificity for the results of Gram stain, mRDT, biochemical tests, conventional culture, and MALDI-ToF MS. Costs related to the index hospitalisation were assumed to be zero after discharge, and hence lifetime costs were the same as costs at 30 days.

The CEM included a spreadsheet calculation to generate costs and outcomes for patients with each different type of pathogen. The key determinant of costs and outcomes in the calculations is the reduction in time to identification of the pathogen and any resistance mechanisms using mRDT compared with conventional culture and MALDI-ToF MS. Consequently, cost estimation focused on aspects of the care pathway which were most likely to be influenced by diagnostic data on the pathogen.

3. Data
   3.1. Clinical data
        3.1.1. Population characteristics

The analysis considers a population of hospitalised adult patients with suspected BSI and following a Gram stain which was positive for a Gram-positive bacteria, a Gram-negative bacteria, or a fungal pathogen. The age and sex distribution of the cohort was taken from Ohnuma 2023 [1]. This publication examined mortality and the empiric therapy for BSIs in United States (US) hospitals. The publication included 32,100 patients from 183 hospitals. Population characteristics are reported according to pathogen type (gram-positive bacteria, gram-negative bacteria, or fungal), and whether empiric therapy was effective. The median age of patients receiving effective therapy for gram-positive bacteria, gram-negative bacteria, or fungal infections was 63, 69 and 62, respectively. An age of 65 was chosen for the cohort as representative of median ages across the three groups. The mean proportion of women across the different cohorts was 45%. Hence the cohort was assumed to comprise 45% women and 55% men.

3.1.2. Pathogen distribution

The pathogen classes considered in the model and the prevalence of each pathogen within the class of pathogens was derived from hospital data collected during the development of the Cobas^®^ Eplex BCID panels in the base case (Table 1). The data represent the distribution of pathogens in the US. Data on the proportion with a resistance mechanism according to pathogen species were also obtained from literature and are tabulated in Table 1 [2-4]. Where data were missing, an assumption of 1% of BSIs with a resistance mechanism was made. A value of zero was applied for all fungal infections. The relative proportion of Gram-positive, Gram-negative and fungal pathogens was calculated from the same source as 33.33%, 62.63% and 4.04% [1].

Table 1: Pathogen prevalence from US epidemiology dataset

| **Pathogen** | **Prevalence (US base case)** | **Prevalence (UK scenario analysis)** | **Proportion with resistance mechanism** |
| --- | --- | --- | --- |
| **Gram-negative** |  |  |  |
| *Acinetobacter* spp (not *baumannii*) | 1.05% | 0.64% | 1%^†^ |
| *Acinetobacter baumannii* | 0.61% | 0.16% | 10.5% |
| *Bacteroides fragilis* | 2.77% | 1.30% | 0% |
| *Citrobacter* spp (not *freundii* & *koseri*) | 1.31% | 0.13% | 2.8% |
| *Citrobacter freundii* & *koseri* | 0.22% | 1.38% | 2.8% |
| *Cronobacter sakazakii* | 0.04% | 0.01% | 0% |
| *Enterobacter* spp (not *cloacae* or *aerogenes*) | 0.04% | 0.18% | 6.9% |
| *Enterobacter cloacae* complex | 3.53% | 2.29% | 5.2% |
| *Enterobacter aerogenes* (*Klebsiella aerogenes*) | 1.26% | 0.57% | 6.9% |
| *Escherichia coli* | 46.47% | 62.17% | 16.8% |
| *Fusobacterium necrophorum* | 0.44% | 0.12% | 0% |
| *Fusobacterium nucleatum* | 0.28% | 0.15% | 0% |
| *Haemophilus influenzae* | 1.05% | 1.04% | 0% |
| *Klebsiella oxytoca* | 2.40% | 1.73% | 6.7% |
| *Klebsiella pneumoniae* | 13.84% | 10.02% | 17.5% |
| *Klebsiella variicola* | 0.78% | 0.49% | 1%^†^ |
| *Klebsiella quasipneumoniae* | 0.00% | 0.00% | 1%^†^ |
| *Morganella morganii* | 1.11% | 0.68% | 0% |
| *Neisseria meningitidis* | 0.02% | 0.76% | 0% |
| *Proteus* spp (not *mirabilis*) | 0.35% | 0.21% | 0% |
| *Proteus mirabilis* | 6.17% | 4.41% | 4.7% |
| *Pseudomonas aeruginosa* | 5.69% | 5.38% | 2.5% |
| *Salmonella* spp | 0.74% | 1.00% | 5.6% |
| Serratia spp (not marcescens) | 0.02% | 0.15% | 0% |
| Serratia marcescens | 1.39% | 1.35% | 1.2% |
| *Stenotrophomonas maltophilia* | 0.59% | 0.41% | 0% |
| *Enterobacteriaceae* / *Enterobacterales* (not listed above) | 0.11% | 0.19% | 1%^†^ |
| Other gram-negative pathogens | 7.72% | 7.49% | 1%^†^ |
| **Gram-positive** |  |  |  |
| *Bacillus subtilis* | 0.48% | 0.49% | 1%^†^ |
| *Bacillus cereus* | 0.26% | 0.15% | 1%^†^ |
| *Corynebacterium* spp | 3.80% | 1.43% | 1%^†^ |
| *Cutibacterium acnes* (*Propionibacterium acnes*) | 1.65% | 1.17% | 1%^†^ |
| *Enterococcus* spp (not *faecalis, faecium, raffinosus*) | 0.25% | 0.50% | 1%^†^ |
| *Enterococcus faecalis* | 3.62% | 3.30% | 4.5% |
| *Enterococcus faecium* | 0.87% | 2.57% | 10.5% |
| *Enterococcus raffinosus* | 0.06% | 0.02% | 1%^†^ |
| *Lactobacillus* spp | 0.44% | 0.27% | 1%^†^ |
| *Listeria* (not *monocytogenes*) | 0.00% | 0.00% | 1%^†^ |
| *Listeria monocytogenes* | 0.03% | 0.15% | 1%^†^ |
| *Micrococcus* spp | 3.22% | 2.34% | 1%^†^ |
| *Staphylococcus aureus* | 15.79% | 14.13% | 41.2% |
| *Staphylococcus* spp, coagulase negative (not speciated) | 4.93% | 47.62% | 1%^†^ |
| *Staphylococcus* spp (speciated other than those listed below) | 2.28% | 0.37% | 1%^†^ |
| *Staphylococcus epidermidis* | 21.68% | 0.00% | 1%^†^ |
| *Staphylococcus lugdunensis* | 0.46% | 0.00% | 1%^†^ |
| *Staphylococcus auricularis* | 0.09% | 0.00% | 1%^†^ |
| *Staphylococcus capitis* | 3.87% | 0.00% | 1%^†^ |
| *Staphylococcus carnosus* | 0.00% | 0.00% | 1%^†^ |
| *Staphylococcus haemolyticus* | 1.44% | 0.00% | 1%^†^ |
| *Staphylococcus hominis* | 11.30% | 0.00% | 1%^†^ |
| *Staphylococcus lentus* | 0.01% | 0.00% | 1%^†^ |
| *Staphylococcus pettenkoferi* | 1.20% | 0.39% | 1%^†^ |
| *Staphylococcus pseudointermedius* | 0.01% | 0.00% | 1%^†^ |
| *Staphylococcus schleiferi* | 0.03% | 0.00% | 1%^†^ |
| *Staphylococcus sciuri* | 0.05% | 0.00% | 1%^†^ |
| *Staphylococcus warneri* | 0.41% | 0.00% | 1%^†^ |
| *Streptococcus* spp (not speciated) | 6.15% | 0.34% | 1%^†^ |
| *Streptococcus* spp (speciated, other than those listed below) | 1.89% | 3.58% | 1%^†^ |
| *Streptococcus agalactiae* (Group B Strep) | 2.45% | 2.92% | 1%^†^ |
| *Streptococcus anginosus* group (*anginosus*, *constellatus* and *intermedius*) | 1.18% | 1.26% | 1%^†^ |
| *Streptococcus gallolyticus* | 0.12% | 0.00% | 1%^†^ |
| *Streptococcus mitis* | 1.05% | 1.09% | 1%^†^ |
| *Streptococcus oralis* | 0.03% | 0.77% | 1%^†^ |
| *Streptococcus pneumoniae* | 2.45% | 6.84% | 1%^†^ |
| *Streptococcus pyogenes* (Group A Strep) | 1.72% | 3.75% | 1%^†^ |
| Other gram-positive pathogens | 4.74% | 4.54% | 1%^†^ |
| **Fungal** |  |  |  |
| *Candida albicans* | 33.20% | 42.97% | 0% |
| *Candida auris* | 0.81% | 0.00% | 0% |
| *Candida dubliniensis* | 2.83% | 2.59% | 0% |
| *Candida famata* | 0.00% | 0.00% | 0% |
| *Candida glabrata* | 36.44% | 30.87% | 0% |
| *Candida guilliermondii* | 0.40% | 0.00% | 0% |
| *Candida kefyr* | 0.00% | 0.16% | 0% |
| *Candida krusei* | 0.40% | 0.00% | 0% |
| *Candida lusitaniae* | 0.81% | 0.86% | 0% |
| *Candida parapsilosis* | 12.96% | 11.78% | 0% |
| *Candida tropicalis* | 5.67% | 3.38% | 0% |
| *Cryptococcus gattii* | 0.00% | 0.39% | 0% |
| *Cryptococcus neoformans* | 1.21% | 1.41% | 0% |
| *Fusarium* | 0.00% | 0.47% | 0% |
| *Rhodotorula* spp | 0.40% | 0.47% | 0% |
| Other fungi | 4.86% | 4.63% | 0% |

†Assumption in place of missing data.
Abbreviations: spp, several species; UK, United Kingdom; US, United States.

For the UK base scenario analysis, data on pathogen prevalence in the United Kingdom (UK) was taken from Public Health England [5]. The data are summarised in (Table 1). The relative proportion of Gram-positive, Gram-negative and fungal pathogens, derived from the Public Health England data were 41.67%, 57.34% and 1.00%, respectively. Data on the proportion of each pathogen with a resistance mechanism were not available for the UK and so these values were unchanged in the scenario analysis.

3.1.3. Pathogen detection across mRDT

Data on pathogens detected by each of the mRDT in the analysis were taken from technical data published by each of the manufacturers [6-9]. The level of information provided on each pathogen varies by pathogen and across mRDTs. Not all pathogen species are uniquely identified. For some pathogens, only the genus is identified; this is identified as a genus call in Table 2. Other species are identified but grouped together. (An example is the *Klebsiella pneumoniae* group call on the Cobas Eplex BCID panels with includes *Klebsiella variicola*, *Klebsiella quasipneumoniae* and *Klebsiella pneumoniae*. These are identified as group calls in Table 2. In the base case, no distinction was made between species, group and genus calls; all were assumed to provide sufficient information to allow assessment of the appropriateness of empiric therapy. In scenario analysis, genus calls or genus and group calls were considered insufficient to allow assessment of the appropriateness of empiric therapy, and treated as if the mRDT had not identified the pathogen.

Table 2: Pathogens detected by each mRDT

| **Pathogen** | **Cobas Eplex BCID panels** | **BioFire BCID panel** | **BioFire BCID2 panel** | **Diasorin Verigene panels** | **Accelerate PhenoTest BC kit** |
| --- | --- | --- | --- | --- | --- |
| **Gram-negative** |  |  |  |  |  |
| *Acinetobacter* spp (not *baumannii*) | No | No | No | Genus | No |
| *Acinetobacter baumannii* | Yes | Yes | Yes | Genus | Yes |
| *Bacteroides fragilis* | Yes | No | Yes | No | No |
| *Citrobacter* spp (not *freundii* & *koseri*) | Genus | No | Genus | Genus | Genus |
| *Citrobacter* spp (*freundii* & *koseri*) | Genus | No | Genus | Genus | Genus |
| *Cronobacter sakazakii* | Yes | No | No | No | No |
| *Enterobacter* spp (not *cloacae* or *aerogenes*) | Group | Genus | Group | Genus | Genus |
| *Enterobacter cloacae* complex | Yes | Yes | Yes | Genus | Genus |
| *Enterobacter aerogenes* (*Klebsiella aerogenes*) | Genus | No | Yes | No | No |
| *Escherichia coli* | Yes | Yes | Yes | Yes | Yes |
| *Fusobacterium necrophorum* | Yes | No | No | No | No |
| *Fusobacterium nucleatum* | Yes | No | No | No | No |
| *Haemophilus influenzae* | Yes | Yes | Yes | No | No |
| *Klebsiella oxytoca* | Yes | Yes | Yes | Yes | Genus |
| *Klebsiella pneumoniae* | Group | Yes | Group | Yes | Group |
| *Klebsiella variicola* | Group | No | No | No | Group |
| *Klebsiella quasipneumoniae* | Group | No | Group | No | Group |
| *Morganella morganii* | Yes | No | Genus | No | No |
| *Neisseria meningitidis* | Yes | Yes | Yes | No | No |
| *Proteus* spp (not *mirabilis*) | Genus | Genus | Genus | Genus | Genus |
| *Proteus mirabilis* | Yes | Genus | Genus | Genus | Genus |
| *Pseudomonas aeruginosa* | Yes | Yes | Yes | Yes | Yes |
| *Salmonella* spp | Genus | No | Genus | Genus | Genus |
| *Serratia* spp (not *marcescens*) | Genus | No | No | No | No |
| *Serratia marcescens* | Yes | Yes | Yes | No | Yes |
| *Stenotrophomonas maltophilia* | Yes | No | Yes | No | No |
| *Enterobacteriaceae* / *Enterobacterales* (not listed above) | No | Genus | Genus | No | No |
| Other gram-negative | No | No | No | No | No |
| **Gram-positive** |  |  |  |  |  |
| *Bacillus subtilis* | Yes | No | No | No | No |
| *Bacillus cereus* | Yes | No | No | No | No |
| *Corynebacterium* spp | Yes | No | No | No | No |
| *Cutibacterium acnes* (*P. acnes*) | Yes | No | No | No | No |
| *Enterococcus* spp (not f*aecalis, faecium, raffinosus*) | Genus | Genus | No | No | No |
| *Enterococcus faecalis* | Yes | Genus | Yes | Yes | Yes |
| *Enterococcus faecium* | Yes | Genus | Yes | Yes | Yes |
| *Enterococcus raffinosus* | Genus | No | No | No | No |
| *Lactobacillus* spp | Genus | No | No | No | No |
| *Listeria* (not *monocytogenes*) | Genus | No | No | Genus | No |
| *Listeria monocytogenes* | Yes | Yes | Yes | No | No |
| *Micrococcus* spp | Genus | No | No | No | No |
| *Staphylococcus aureus* | Yes | Yes | Yes | Yes | Yes |
| *Staphylococcus* spp, coagulase negative (not speciated) | Genus | Genus | Genus | Genus | No |
| *Staphylococcus* spp (speciated other than those listed below) | Genus | Genus | Genus | Genus | No |
| *Staphylococcus epidermidis* | Yes | Genus | Yes | Yes | Genus |
| *Staphylococcus lugdunensis* | Yes | Genus | Yes | Yes | Yes |
| *Staphylococcus auricularis* | Genus | No | Genus | Genus | No |
| *Staphylococcus capitis* | Genus | Genus | Genus | Genus | Genus |
| *Staphylococcus carnosus* | Genus | No | Genus | Genus | No |
| *Staphylococcus haemolyticus* | Genus | Genus | Genus | Genus | Genus |
| *Staphylococcus hominis* | Genus | Genus | Genus | Genus | Genus |
| *Staphylococcus lentus* | Genus | No | No | Genus | No |
| *Staphylococcus pettenkoferi* | Genus | No | Genus | Genus | No |
| *Staphylococcus pseudointermedius* | Genus | No | Genus | Genus | No |
| *Staphylococcus schleiferi* | Genus | No | Genus | Genus | No |
| *Staphylococcus sciuri* | Genus | No | Genus | Genus | No |
| *Staphylococcus warneri* | Genus | Genus | Genus | Genus | Genus |
| *Streptococcus* spp (not speciated) | Genus | Genus | Genus | Genus | Genus |
| *Streptococcus* spp (speciated, other than those listed below) | Genus | Genus | Genus | Genus | Genus |
| *Streptococcus agalactiae* (Group B Strep) | Yes | Yes | Yes | Yes | No |
| *Streptococcus anginosus* group (*anginosus, constellatus and intermedius*) | Yes | Genus | Genus | Yes | No |
| *Streptococcus gallolyticus* | Genus | Genus | Genus | Genus | Genus |
| *Streptococcus mitis* | Genus | Genus | Genus | Genus | Genus |
| *Streptococcus oralis* | Genus | Genus | Genus | Genus | Genus |
| *Streptococcus pneumoniae* | Yes | Yes | Yes | Yes | No |
| *Streptococcus pyogenes* (Group A Strep) | Yes | Yes | Yes | Yes | No |
| Other gram-positive | No | No | No | No | No |
| **Fungal** |  |  |  |  |  |
| *Candida albicans* | Yes | Yes | Yes | No | Yes |
| *Candida auris* | Yes | No | Yes | No | No |
| *Candida dubliniensis* | Yes | No | No | No | No |
| *Candida famata* | Yes | No | No | No | No |
| *Candida glabrata* | Yes | Yes | Yes | No | Yes |
| *Candida guilliermondii* | Yes | No | No | No | No |
| *Candida kefyr* | Yes | No | No | No | No |
| *Candida krusei* | Yes | Yes | Yes | No | No |
| *Candida lusitaniae* | Yes | No | No | No | No |
| *Candida parapsilosis* | Yes | Yes | Yes | No | No |
| *Candida tropicalis* | Yes | Yes | Yes | No | No |
| *Cryptococcus gattii* | Yes | No | Group | No | No |
| *Cryptococcus neoformans* | Yes | No | Group | No | No |
| *Fusarium* | Yes | No | No | No | No |
| *Rhodotorula* spp | Yes | No | No | No | No |
| Other fungi | No | No | No | No | No |

†Assumption in place of missing data.
Abbreviations: spp, several species.

3.1.4. Composition and effectiveness of empiric antimicrobial therapy

Data on the composition and effectiveness of antimicrobial therapy was taken from Figure 2 in Ohnuma 2023 [1] which was digitised to extract the data on effectiveness of empiric therapy by pathogen class. The most appropriate value was then applied to each of the 82 pathogens in the model (Table 3).

Table 3: Effectiveness of empiric therapy by type of pathogen

| **Pathogen** | **Effectiveness** |
| --- | --- |
| **Gram-negative** |  |
| *Acinetobacter* spp | 64.6% |
| *Bacteroides fragilis* | 90.4% |
| *Citrobacter* spp | 89.2% |
| *Cronobacter sakazakii* | 90.4% |
| *Enterobacter* spp | 93.2% |
| *Escherichia coli* | 95.7% |
| *Fusobacterium necrophorum* | 90.4% |
| *Fusobacterium nucleatum* | 90.4% |
| *Haemophilus influenzae* | 90.4% |
| *Klebsiella* spp | 95.6% |
| *Morganella morganii* | 90.4% |
| *Neisseria meningitidis* | 90.4% |
| *Proteus* spp | 96.2% |
| *Pseudomonas aeruginosa* | 88.6% |
| *Salmonella* spp | 90.4% |
| *Serratia* spp | 93.8% |
| *Stenotrophomonas maltophilia* | 90.4% |
| *Enterobacteriaceae* / *Enterobacterales* (not listed above) | 93.2% |
| Other gram-negative | 90.4% |
| **Gram-positive** |  |
| *Bacillus* spp | 95.1% |
| *Corynebacterium* spp | 95.1% |
| *Cutibacterium acnes* (*P. acnes*) | 95.1% |
| *Enterococcus* spp | 91.4% |
| *Lactobacillus* spp | 95.1% |
| *Listeria* spp | 95.1% |
| *Micrococcus* spp | 95.1% |
| *Staphylococcus* spp | 98.2% |
| *Streptococcus* spp | 96.4% |
| Other gram-positive | 95.1% |
| **Fungal** |  |
| All fungi | 65.1% |

Abbreviations: spp, several species.

The composition of empiric therapy was also reported for patients according to pathogen class and whether empiric therapy was effective (Table 4). The data indicate a mean number of antimicrobials in empiric therapy of 2.24, 2.30 and 2.83 for patients with a Gram-negative, Gram-positive, and fungal infection, respectively. The data appear consistent with empiric therapy consisting of two broad-spectrum antibiotics active against Gram-positive and Gram-negative bacteria, with the addition of a fungal agent in patients suspected to have a fungal infection. The data were used to calculate days of therapy with respect to *C. difficile* infection risk and the cost of empiric therapy.

Table 4: Composition of empiric therapy pathogen class and effectiveness of empiric therapy

|  | **Gram-negative** | | **Gram-positive** | | **Candida** | |
| --- | --- | --- | --- | --- | --- | --- |
| **Medication** | **Ineffective (n=841)** | **Effective (n=14,114)** | **Ineffective (n=512)** | **Effective (n=16,341)** | **Ineffective (n=102)** | **Effective (n=190)** |
| Cephalosporin | 455 (54.1%) | 9,008 (63.8%) | 287 (56.1%) | 10,334 (63.2%) | 52 (51.0%) | 116 (61.1%) |
| Penicillin | 347 (41.3%) | 7,478 (53.0%) | 184 (35.9%) | 8,657 (53.0%) | 59 (57.8%) | 90 (47.4%) |
| Carbapenem | 144 (17.1%) | 4,291 (30.4%) | 42 (8.2%) | 1,787 (10.9%) | 15 (14.7%) | 47 (24.7%) |
| Quinolone | 104 (12.4%) | 2,330 (16.5%) | 75 (14.6%) | 1,825 (11.2%) | 11 (10.8%) | 22 (11.6%) |
| Vancomycin | 505 (60.0%) | 8,053 (57.1%) | 178 (34.8%) | 14,772 (90.4%) | 68 (66.7%) | 148 (77.9%) |
| Antifungal | 46 (5.5%) | 687 (4.9%) | 21 (4.1%) | 566 (3.5%) | 8 (7.8%) | 190 (100%) |

3.1.5. Proportion of patients optimised from effective broad-spectrum antimicrobial therapy to targeted therapy

Data on optimisation from effective broad-spectrum therapy was piecemeal and diverse with reported rates varying from 12.5% to 98% [10]. The proportion of patients optimised (de-escalated) from effective broad-spectrum antimicrobial therapy to a targeted therapy following identification of the pathogen or resistance mechanism was taken from a US study reporting data for a tertiary care medical centre [11]. The study reported 62.9% of patients optimised at three days.

3.1.6. Mortality

Thirty-day mortality following BSI has been reported as a function of the infectious species using a population wide sample of 22,935 BSI in 19,326 patients in Ontario in 2017 [12]. Over 80% of the patients with a BSI were aged 50 years and over and 45% were female. The overall mortality rate was 17.0%. The highest mortality rate of 41.9% was observed for *Clostridiodes* species. The data in the study were matched to the model where possible. Mortality rates were not reported for the gram-negative species *Cronobacter sakazakii, Morganella morganii, Neisseria meningitidis,* and *Salmonella* several species (spp). A mean mortality across the remaining gram-negative species of 18.45% was applied for these species. Mortality rates were not reported for the gram-positive species *Cutibacterium acnes, Lactobacillus* spp, *Listeria* spp and *Micrococcus* spp. A mean mortality across the remaining gram-positive species of 18.85% was applied for these species. The mortality rate by type of pathogen is tabulated in Table 5.

Table 5: Thirty-day Pathogen mortality

| **Pathogen** | **Mortality** | **Pathogen species in source data** |
| --- | --- | --- |
| **Gram-negative** |  |  |
| *Acinetobacter* spp (non-*baumannii*) | 15.5% | *Acinetobacter* spp |
| *Acinetobacter baumannii* | 15.5% | *Acinetobacter* spp |
| *Bacteroides fragilis* | 25.3% | *Bacteroides fragilis* |
| *Citrobacter* spp (not *freundii* & *koseri*) | 15.6% | *Citrobacter* spp |
| *Citrobacter* spp (*freundii* & *koseri*) | 15.6% | *Citrobacter* spp |
| *Cronobacter sakazakii* | 18.5% | N/A |
| *Enterobacter* spp (not *cloacae* or *aerogenes*) | 19.8% | *Enterobacter* spp |
| *Enterobacter cloacae* complex | 19.8% | *Enterobacter* spp |
| *Enterobacter aerogenes* (*Klebsiella aerogenes*) | 19.8% | *Enterobacter* spp |
| *Escherichia coli* | 12.1% | *Escherichia coli* |
| *Fusobacterium necrophorum* | 13.0% | *Fusobacterium* spp |
| *Fusobacterium nucleatum* | 13.0% | *Fusobacterium* spp |
| *Haemophilus influenzae* | 19.5% | *Haemophilus influenzae* |
| *Klebsiella oxytoca* | 17.6% | *Klebsiella* spp |
| *Klebsiella pneumoniae* | 17.6% | *Klebsiella* spp |
| *Klebsiella variicola* | 17.6% | *Klebsiella* spp |
| *Klebsiella quasipneumoniae* | 17.6% | *Klebsiella* spp |
| *Morganella morganii* | 18.5% | N/A |
| *Neisseria meningitidis* | 18.5% | N/A |
| *Proteus* spp (not *mirabilis*) | 20.7% | *Proteus* spp |
| *Proteus mirabilis* | 20.7% | *Proteus* spp |
| *Pseudomonas aeruginosa* | 24.7% | *Pseudomonas* spp |
| *Salmonella* spp | 18.5% | N/A |
| *Serratia* spp (not marcescens) | 20.7% | *Serratia* spp |
| *Serratia marcescens* | 20.7% | *Serratia* spp |
| *Stenotrophomonas maltophilia* | 23.2% | *Stenotrophomonas maltophilia* |
| *Enterobacteriaceae* / *Enterobacterales* (not listed above) | 19.8% | *Enterobacter* spp |
| Other gram-negative | 18.5% | N/A |
| **Gram-positive** |  |  |
| *Bacillus subtilis* | 0% (13.1%) | *Bacillus* spp† |
| *Bacillus cereus* | 0% (13.1%) | *Bacillus* spp† |
| *Corynebacterium* spp | 0% (27.3%) | *Corynebacterium* spp† |
| *Cutibacterium acnes* (*P. acnes*) | 0% (18.9%) | N/A† |
| *Enterococcus* spp (not *faecalis, faecium, raffinosus*) | 23.6% | *Enterococcus* spp |
| *Enterococcus faecalis* | 23.6% | *Enterococcus spp* |
| *Enterococcus faecium* | 23.6% | *Enterococcus* spp |
| *Enterococcus raffinosus* | 23.6% | *Enterococcus* spp |
| *Lactobacillus* spp | 0% (18.9%) | N/A† |
| *Listeria* (not *monocytogenes*) | 18.9% | N/A |
| *Listeria monocytogenes* | 18.9% | N/A |
| *Micrococcus* spp | 0% (18.9%) | N/A† |
| *Staphylococcus aureus* | 22.8% | *Staphylococcus aureus* |
| *Staphylococcus* spp, coagulase negative (not speciated) | 0% (19.7%) | Other coagulase negative *Staphylococcus*† |
| *Staphylococcus* spp (speciated other than those listed below) | 0% (19.7%) | Other coagulase negative *Staphylococcus*† |
| *Staphylococcus epidermidis* | 0% (19.7%) | Other coagulase negative *Staphylococcus*† |
| *Staphylococcus lugdunensis* | 17.6% | *Staphylococcus lugdunensis* |
| *Staphylococcus auricularis* | 19.7% | Other coagulase negative *Staphylococcus*† |
| *Staphylococcus capitis* | 0% (19.7%) | Other coagulase negative *Staphylococcus*† |
| *Staphylococcus carnosus* | 0% (19.7%) | Other coagulase negative *Staphylococcus*† |
| *Staphylococcus haemolyticus* | 0% (19.7%) | Other coagulase negative *Staphylococcus*† |
| *Staphylococcus hominis* | 0% (19.7%) | Other coagulase negative *Staphylococcus*† |
| Staphylococcus lentus | 0% (19.7%) | Other coagulase negative *Staphylococcus*† |
| *Staphylococcus pettenkoferi* | 0% (19.7%) | Other coagulase negative *Staphylococcus*† |
| *Staphylococcus pseudointermedius* | 0% (19.7%) | Other coagulase negative *Staphylococcus*† |
| *Staphylococcus schleiferi* | 0% (19.7%) | Other coagulase negative *Staphylococcus*† |
| *Staphylococcus sciuri* | 0% (19.7%) | Other coagulase negative *Staphylococcus*† |
| *Staphylococcus warneri* | 0% (19.7%) | Other coagulase negative *Staphylococcus*† |
| *Streptococcus* spp (not speciated) | 0% (16.7%) | Other *Streptococcus* species† |
| *Streptococcus* spp (speciated, other than those listed below) | 0% (16.7%) | Other *Streptococcus* species† |
| *Streptococcus agalactiae* (Group B Strep) | 13.8% | *Streptococcus agalactiae* |
| *Streptococcus anginosus* group (a*nginosus, constellatus* and *intermedius*) | 14.2% | Viridans group *Streptococcus* |
| *Streptococcus gallolyticus* | 16.7% | Other *Streptococcus* species† |
| *Streptococcus mitis* | 13.0% | *Streptococcus mitis* |
| *Streptococcus oralis* | 16.7% | Other *Streptococcus* species† |
| *Streptococcus pneumoniae* | 15.4% | *Streptococcus pneumoniae* |
| Streptococcus pyogenes (Group A Strep) | 15.8% | *Streptococcus pyogenes* |
| Other gram-positive | 18.9% | N/A |
| **Fungal** |  |  |
| All fungal | 32.0% | *Candida* spp |

†Species assumed to be a contaminant in the base case and hence associated mortality assumed to be zero in the base case.
Abbreviations: N/A, not applicable; spp, several species.

3.1.7. Relationship between time to effective therapy and mortality

The relationship between time to effective therapy and mortality has been noted in a number of studies, albeit much of the existing literature consists of small studies [13, 14]. Recently, data linking time to effective therapy and mortality have been reported using a relatively large representative patient sample with granularity of analysis over delays from 1–‍72 hours [15]. The Swedish study examined 10,328 BSI in 9,192 patients over the period 2012-2019. The data show a clear dose-response relationship with mortality increasing with time to effective therapy, albeit a statistically significant increase in mortality (at the 5% level) was not observed until a delay of 12 hours. The reported odds ratios (OR) at 1, 3, 6, 12, 24, 48, and 72 hours were modelled using linear regression, which generated a slope of 0.010 hour^-1^ and an intercept of 0.958. The reported ORs and the ORs predicted by the linear regression are reported in Table 6. The raw data and the modelled values are plotted in Figure 3. It is evident that the relationship between the OR of death and time to effective therapy is broadly linear over the period of 3–72 hours. The use of a linear function allows an estimation of the impact of each hour of delay which is invariant to the baseline time to effective therapy.

Table 6: ORs as a function of time to effective therapy, raw data, and modelled values

| **Time to effective therapy** | **Reported OR (95% CI)** | **OR from fitted regression** |
| --- | --- | --- |
| 1 hour | 0.83 (0.72, 0.95) | 0.97 |
| 3 hours | 1.00 (0.84, 1.15) | 0.99 |
| 6 hours | 1.05 (0.91, 1.22) | 1.02 |
| 12 hours | 1.17 (1.01, 1.37) | 1.08 |
| 24 hours | 1.24 (1.04, 1.47) | 1.20 |
| 48 hours | 1.41 (1.15, 1.74) | 1.44 |
| 72 hours | 1.67 (1.30, 2.15) | 1.68 |

Abbreviations: CI, confidence interval; OR, odds ratio.

Figure 3: Reported and modelled OR of mortality as a function of time to effective therapy

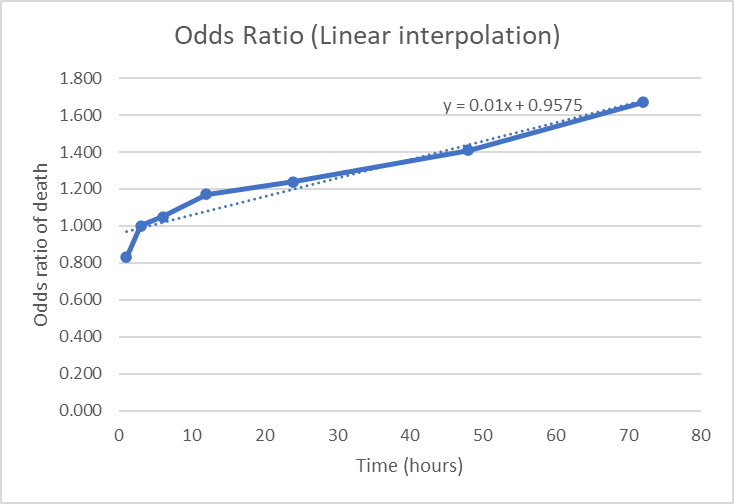

Abbreviations: OR, odds ratio.

3.1.8. Time to diagnosis with MALDI-ToF MS and mRDT

The studies reporting data on time to diagnosis with conventional culture and MALDI-ToF MS were of variable quality and the data reported was disparate [16-19]. The time was assumed to be 24 hours (after Gram stain) for non-resistant pathogens and 48 hours for resistant pathogens on the basis of the requirement for overnight culturation for conventional MALDI-ToF MS and a further overnight culturation for susceptibility testing. The time is assumed to include the time taken to inform the clinician treating the patient and for the clinician to initiate any changes to the patients’ therapy. The corresponding time for diagnosis with mRDT was assumed to be 3 hours.

3.1.9. Risk of *C. difficile* Infection and associated mortality

The risk of *C. difficile* infection is generally accepted to be a function of the duration of antimicrobial therapy [20]. Data on *C. difficile* infections were taken from a US study on the cumulative impact of antibiotic exposure on the risk of *C. difficile* infection [21]. The study reports data on 10,154 hospitalisations in 7,792 patients at the Strong Memorial Hospital in New York in 2005. Patients included in the study were aged 18 years and older and had received antibiotics for at least two days. In total, 241 cases of *C. difficile* were observed. Data are presented as a function of defined daily doses of antibiotics – total dose of each antibiotic received was converted to the number of daily doses according to the World Health Organization Defined Daily Dose system, and then the totals summed across concomitant therapies. Data from the study on *C. difficile* infections by category of defined daily dose is reported in Table 7.

Table 7: Crude rates of *C. difficile* infection as a function of defined daily doses of antibiotics

| **Defined daily dose** | **Patients with *C. difficile* infection** | **Patients without *C. difficile* infection** | **Probability of *C. difficile* infection** |
| --- | --- | --- | --- |
| <3.0 | 18 | 1502 | 1.20% |
| 3.0 to 7.79 | 49 | 3702 | 1.32% |
| 7.8 to 21.0 | 89 | 2952 | 3.01% |
| >21.0 | 85 | 1757 | 4.84% |

An assumption was made that the risk of *C. difficile* infection is linear between the midpoints of the two middle bands (3.0 to 7.79, and 7.8 to 21.0), and that the value reported for the two middle bands is representative of the crude probability of infection at the midpoint values of 5.4 and 14.4 defined daily doses. The gradient of the line joining the two points is 0.19% per defined daily dose. The CEM assumed that the risk of *C. difficile* increased linearly at a rate of 0.19% per defined daily dose of antibiotic.

Mortality associated with *C. difficile* infection varies according to the associated comorbidities of the patient sample. A recent review of studies reported 30-day mortality rates varying from 8% to 53% [22]. Another recent US study utilised data from the National Hospital Discharge Survey (NHDS) on 162 million patients and identified 1.26 million cases of *C. difficile* [23]. A crude mortality rate of 6.9% for patients with *C. difficile* was reported, and an OR of mortality of 1.45 associated with *C. difficile* infection. Notably, this study was not restricted to patients with BSI. Data on mortality for patients with a BSI and an associated *C. difficile* infection is limited. Data were available for a population of patients with sepsis and *C. difficile* infection [24]. The authors undertook a propensity-scored matched analysis to report an absolute increased risk of 8.6% (95% confidence interval [CI]: 6.4, 10.9), and a relative risk of 1.6 (95% CI: 1.4, 1.8) associated with *C. difficile* infection. The model assumed an absolute risk of death of 8.6% associated with *C. difficile* infection.

3.1.10. Risk of AKI and associated mortality

The risk of AKI with vancomycin is well-documented [25]. However, other antibiotics are also associated with a risk of AKI [26]. The CEM assumed a risk of AKI whilst patients remained on broad-spectrum empiric therapy without differentiating the impact of different components of empiric therapy. It should be noted that the available data on empiric therapy indicated that the majority of patients receive vancomycin, and this rises to 90% of patients who received effective empiric therapy for a gram-positive BSI (Table 4). Data on the risk of AKI were taken from a US study of the risk of AKI which compared patients receiving ≥4 g/day of vancomycin with patients receiving <4 g/day of vancomycin and patients receiving linezolid [27]. Patients were included if they were aged over 18 years, non-neutropenic and in receipt of therapy for >2 days. The study included 220 patients receiving <4 g/day of vancomycin. The Kaplan-Meier (KM) for time to AKI was digitised. The plot is insufficiently granular to allow extraction of the pseudo-patient data. Survival without nephrotoxicity was 83.3% at day 12 for patients receiving <4 g/day of vancomycin. Beyond this point the curve plateaus. The risk of AKI was assumed to be constant over time with exposure to empiric antimicrobials up to day 12. A daily risk of 1.51% was applied in the CEM. This equates to an AKI-free survival of 83.3% at 12 days.

The impact of AKI on mortality was estimated from a large recent US study examining the incidence and outcome of AKI in patients with carbapenem-resistant gram-negative infections [28]. The study included 750 patients, of whom 91 were diagnosed with AKI. Unadjusted in-hospital mortality was 5.3% and 20.9% and 5.3% in patients with and without AKI. After adjustment for differences in patient characteristics across the two groups using inverse probability weighting, in-hospital mortality was 18.5% and 5.6% for patients with and without AKI. The absolute difference in in-hospital mortality from the adjusted analysis of 12.9% was assumed as the mortality attributable to AKI. It should be noted that in-hospital mortality was modestly higher than 30-day mortality, reflecting stays longer than 30 days for some patients who ultimately died and associated outcomes using the acute kidney injury network (AKIN) criteria to define AKI for those who died before discharge.

3.1.11. LOS in hospital

Data on LOS with a BSI in the base case was taken from a study of the impact of ineffective empiric antimicrobial therapy on LOS undertaken at the Palmetto Health Richland and Baptist Hospitals in Columbia between 2010 and 2013 [29]. The study recruited 830 patients with a gram-negative BSI, of which 72 (9%) received ineffective empiric therapy. Median LOS for patients receiving effective empiric therapy was not reported, but data were plotted as the KM curve over 30 days. The data were digitised and the area under the KM curve for patients receiving effective antimicrobial therapy was estimated as 11.56 days. This restricted mean value was applied in the CEM for patients receiving appropriate empiric therapy.

Data on LOS in the UK was taken from a UK study of 273,001 patients with sepsis which reported a median LOS of 14 days [30].

3.1.12. Impact of delay on time to effective therapy on LOS

Data on the impact of time to effective therapy on LOS were limited. The CEM exploited data from a US study examining the impact of time to effective therapy on LOS [31]. The study recruited 917 patients aged 16 years and over with a BSI between 1999 and 2000. The study reported a duration ratio of 1.004 (95% CI: 1.001, 1.008) for each hour of delay to Gram stain notification. A duration ratio of 1.004 per hour indicates that for every additional hour of delay to Gram stain notification, the patient’s LOS is expected to be multiplied by 1.004. In practical terms, each extra hour of delay is associated with about a 0.4% increase in the LOS. The duration ratio was applied in the model to calculate the impact of each hour of delay to effective therapy.

3.1.13. Impact of *C. difficile* and AKI on LOS

The impact of *C. difficile* infection on LOS was taken from a recent systematic review which grouped studies according to the robustness of the methodology used to estimate increased LOS [32]. The more robust methodologies of time-varying matching, multistate modelling and survival analysis generate mean additional LOS for *C. difficile* infection of 3.66 (95% CI: 0.92, 6.40), 2.32 (95% CI: 0.97, 3.68), and 6.00 (95% CI: 2.43, 9.57) days, respectively. The estimate for survival analysis utilised a single study. The estimate for time-varying matched analyses was selected (3.66 days) as the estimate pooled observed data from three studies.

Data on the impact of AKI on LOS was taken from the study which provided data on the impact of AKI on mortality (Section 3.1.10) [33]. The study reported a median increase in total LOS of 4 days for patients with an AKI compared with those without. The median increase in LOS after vancomycin initiation was also 4 days for patients with an AKI compared with those without. The median LOS for patients with and without AKI were 11 (interquartile range [IQR]: 8, 17) and 15 (IQR: 11, 23), respectively. The median LOS were converted to mean LOS of 11.53 and 15.84 days using the method of Wan et al. [34]. The resulting difference between the mean values of 4.31 days was applied in the CEM as the absolute increase in LOS for patient experiencing AKI.

3.2. Cost data

The following cost data were included in the economic evaluation:

- Test costs for mRDT and MALDI-ToF MS
- Cost of empiric and targeted antimicrobials
- Cost of hospitalisation (general ward).

The CEM did not include productivity costs or any costs following discharge from hospital (either related or unrelated to the BSI episode). The model assumed that patients surviving to 30 days are discharged from hospital and make a full recovery. Costs of conventional culture, Gram stain, biochemical tests (e.g. catalase test, oxidate test, coagulase test, Triple Sugar Iron [TSI] agar test) and AST were excluded. These costs accrue in both the standard of care approach and strategies including mRDT. As a result, these costs do not impact incremental costs. Costs of an ASP were also excluded on the assumption that such a programme is already in place and any costs would not be impacted by adoption of mRDT. Costs associated with AKI or *C. difficile* other than increased LOS on a general ward were also not included. Potential costs of AKI and *C. difficile* that were not considered include costs of renal replacement therapy and any requirement for critical care.

3.2.1. Cost of mRDT and MALDI-ToF MS

Costs for mRDT were obtained from internal evidence generation undertaken by Roche. The costs applied in the base case are shown in Table 8.

Table 8: Cost of mRDT

| **Test** | **Cost** |
| --- | --- |
| Cobas Eplex BCID panels | $115 |
| BioFire BCID panel | $110 |
| BioFire BCID2 panel | $115 |
| Diasorin Verigene BCID panels | $64 |
| Accelerate PhenoTest BC kit | $168 |

Abbreviations: BCID, blood culture identification; mRDT, molecular rapid diagnostic test.

Patients receiving mRDT were also assumed to undergo testing with conventional culture and MALDI-ToF MS. Hence the cost of MALDI-ToF MS is the same across all comparators and cancels out. However, the cost was still estimated for completeness. The purchase cost of a MALDI-ToF mass spectrometer was taken as $270,000 as reported in cost study of MALDI-ToF MS [35]. Assuming a five-year lifespan, the annual cost is $58,996 after amortization at 3%. The cost study of MALDI-ToF MS also reports reagent costs of $2.69 per isolate and estimates 21,930 isolates per year. On this basis, the capital cost per isolate is $2.69, and the total cost per isolate, including reagents, is $5.83. The cost was inflated from 2015 to 2024 using the medical care inflation index in the US, city average for all urban consumers, to generate a cost of $7.52 [36].

3.2.2. Costs of empiric and targeted antimicrobials

The specific drugs comprising antimicrobials therapy were identified by the clinical authors (AT and TS) (Table 9).

Table 9: Component drugs assumed for empiric antimicrobial therapy

| **Antimicrobial class** | **Antimicrobial assumed** |
| --- | --- |
| Cephalosporin | Cefepime |
| Penicillin | Ampicillin |
| Carbapenem | Meropenem |
| Quinolone | Levofloxacin |
| Vancomycin | Vancomycin |
| Antifungal | Micafungin |

The most commonly used targeted antimicrobial for each pathogen was also identified by the clinical authors (AT and TS) (Table 10).

Table 10: Targeted antimicrobial assumed for each pathogen species in the model

| **Pathogen** | **Targeted therapy** |
| --- | --- |
| **Gram-negative** |  |
| *Acinetobacter baumannii* | Meropenem |
| *Acinetobacter* spp (non-*baumannii*) | Meropenem |
| *Bacteroides fragilis* | Metronidazole |
| *Citrobacter* spp (not *freundii* & *koseri*) | Cefepime |
| *Citrobacter freundii* & *koseri* | Cefepime |
| *Cronobacter sakazakii* | Cefepime |
| *Enterobacter* spp (not *cloacae* or *aerogenes*) | Cefepime |
| *Enterobacter cloacae* complex | Cefepime |
| *Enterobacter aerogenes* (*Klebsiella aerogenes*) | Cefepime |
| *Escherichia coli* | Ceftriaxone |
| *Fusobacterium necrophorum* | Piperacillin-tazobactam |
| *Fusobacterium nucleatum* | Piperacillin-tazobactam |
| *Haemophilus influenzae* | Ampicillin-sulbactam |
| *Klebsiella oxytoca* | Ceftriaxone |
| *Klebsiella pneumoniae* | Ceftriaxone |
| *Klebsiella variicola* | Ceftriaxone |
| *Klebsiella quasipneumoniae* | Ceftriaxone |
| *Morganella morganii* | Cefepime |
| *Neisseria meningitidis* | Ceftriaxone |
| *Proteus* spp (not *mirabilis*) | Cefepime |
| *Proteus mirabilis* | Ceftriaxone |
| *Pseudomonas aeruginosa* | Cefepime |
| *Salmonella* spp | Ceftriaxone |
| *Serratia* spp (not *marcescens*) | Cefepime |
| *Serratia marcescens* | Cefepime |
| *Stenotrophomonas maltophilia* | Trimethoprim/sulfamethoxazole |
| *Enterobacteriaceae* / *Enterobacterales* (not listed above) | Ceftriaxone |
| Other gram-negative | Cefepime |
| **Gram-positive** |  |
| *Bacillus subtilis* | Stop therapy (vancomycin)† |
| *Bacillus cereus* | Stop therapy (vancomycin)† |
| *Corynebacterium* spp | Stop therapy (vancomycin)† |
| *Cutibacterium acnes* (*P. acnes*) | Stop therapy (penicillin G)† |
| *Enterococcus* spp (not *faecalis, faecium, raffinosus*) | Ampicillin |
| *Enterococcus faecalis* | Ampicillin |
| *Enterococcus faecium* | Daptomycin |
| *Enterococcus raffinosus* | Ampicillin |
| *Lactobacillus* spp | Stop therapy (ampicillin)† |
| *Listeria* (not *monocytogenes*) | Ampicillin + gentamicin |
| *Listeria monocytogenes* | Ampicillin + gentamicin |
| *Micrococcus* spp | Stop therapy (penicillin G)† |
| *Staphylococcus aureus* | Cefazolin |
| *Staphylococcus* spp, coagulase negative (not speciated) | Stop therapy (cefazolin)† |
| *Staphylococcus* spp (speciated other than those listed below) | Stop therapy (cefazolin)† |
| *Staphylococcus epidermidis* | Stop therapy (cefazolin)† |
| *Staphylococcus lugdunensis* | Stop therapy (cefazolin)† |
| *Staphylococcus auricularis* | Stop therapy (cefazolin)† |
| *Staphylococcus capitis* | Stop therapy (cefazolin)† |
| *Staphylococcus carnosus* | Stop therapy (cefazolin)† |
| *Staphylococcus haemolyticus* | Stop therapy (cefazolin)† |
| *Staphylococcus hominis* | Stop therapy (cefazolin)† |
| *Staphylococcus lentus* | Stop therapy (cefazolin)† |
| *Staphylococcus pettenkoferi* | Stop therapy (cefazolin)† |
| *Staphylococcus pseudointermedius* | Stop therapy (cefazolin)† |
| *Staphylococcus schleiferi* | Stop therapy (cefazolin)† |
| *Staphylococcus sciuri* | Stop therapy (cefazolin)† |
| *Staphylococcus warneri* | Stop therapy (cefazolin)† |
| *Streptococcus* spp (not speciated) | Stop therapy (cefazolin)† |
| *Streptococcus* spp (speciated, other than those listed below) | Stop therapy (penicillin G)† |
| *Streptococcus agalactiae* (Group B Strep) | Penicillin G |
| *Streptococcus anginosus* group (*anginosus, constellatus* and *intermedius*) | Penicillin G |
| *Streptococcus gallolyticus* | Penicillin G |
| *Streptococcus mitis* | Penicillin G |
| *Streptococcus oralis* | Penicillin G |
| *Streptococcus pneumoniae* | Ceftriaxone |
| *Streptococcus pyogenes* (Group A Strep) | Penicillin G |
| Other gram-positive | Vancomycin |
| **Fungal** |  |
| *Candida albicans* | Micafungin |
| *Candida auris* | Micafungin |
| *Candida dubliniensis* | Micafungin |
| *Candida famata* | Micafungin |
| *Candida glabrata* | Micafungin |
| *Candida guilliermondii* | Amphotericin B |
| *Candida kefyr* | Micafungin |
| *Candida krusei* | Micafungin |
| *Candida lusitaniae* | Micafungin |
| *Candida parapsilosis* | Micafungin |
| *Candida tropicalis* | Micafungin |
| *Cryptococcus gattii* | Amphotericin B + flucytosine |
| *Cryptococcus neoformans* | Amphotericin B + flucytosine |
| *Fusarium* | Amphotericin B + voriconazole |
| *Rhodotorula* spp | Amphotericin B |
| Other fungi | Amphotericin B |

†Base case assumes species is a contaminant.
Abbreviations: spp, several species.

Unit costs for drugs were sourced from the US Veteran Affairs drug pricing database [37]. Data on dosing was sourced from the British National Formulary (BNF). Dose calculations requiring body mass assumed an average mass of 199.8 lb and 170.8 lb for US men and women reported by the Centre for Disease Control [38]. Costs are summarised in Table 11. Unit costs for drugs for the UK scenario analysis were derived from the BNF [39].

Table 11: Daily costs of antimicrobial therapy

| **Antimicrobial** | **Cost per dose ($) (US base case)** | **Doses per day** | **Cost per day ($) (US base case)** | **Cost per dose (£) (UK scenario analysis)** |
| --- | --- | --- | --- | --- |
| Ampicillin | 2.21 | 6 | 13.25 | 32.88 |
| Ampicillin + gentamicin | 33.49 | 1 | 33.49 | 32.88 |
| Ampicillin-sulbactam | 3.02 | 4 | 12.06 | 32.88 |
| Cefazolin | 1.79 | 3 | 5.37 | 16.18 |
| Cefepime | 19.33 | 3 | 57.99 | 11.00 |
| Cefiderocol | 412.09 | 3 | 1,236.28 | 131.90 |
| Ceftazidime-avibactam | 344.67 | 3 | 1,034.01 | 85.70 |
| Ceftriaxone | 1.31 | 2 | 2.61 | 6.50 |
| Daptomycin | 22.71 | 1 | 22.71 | 88.00 |
| Levofloxacin | 10.67 | 1 | 10.67 | 4.00 |
| Meropenem | 10.06 | 3 | 30.19 | 16.00 |
| Meropenem-vaborbactam | 250.27 | 3 | 750.80 | 55.67 |
| Metronidazole | 1.04 | 3 | 3.11 | 4.81 |
| Penicillin G | 1.92 | 6 | 11.49 | 13.33 |
| Piperacillin-tazobactam | 4.52 | 4 | 18.09 | 4.80 |
| Trimethroprim/sulfamethoxazole | 6.91 | 2 | 13.82 | 4.72 |
| Vancomycin | 5.51 | 3 | 16.52 | 11.25 |
| Amphotericin B | 14.64 | 1 | 14.64 | 164.38 |
| Amphotericin B + 5FC | 191.46 | 1 | 191.46 | 164.38 |
| Amphotericin B + voriconazole | 51.54 | 1 | 51.54 | 164.38 |
| Micafungin | 76.18 | 1 | 76.18 | 341.00 |

Abbreviations: 5FC, flucytosine; UK, United Kingdom; US, United States.

3.2.3. Cost of hospital stay

The costs of hospital stay per inpatient day was taken from data published by the Kaiser Family Foundation (KFF) [40]. The US average value of $3,025 was selected. The cost of a bed day in hospital in the UK of £933 was taken from a study of the costs associated with healthcare -associated infections in the UK [41]. The study reported a cost of £799 (2017/18 Pounds Sterling [GBP]) which was inflated to £933 (2022/23 GBP).

3.3. Data on quality of life and life expectancy

Patients alive at 30 days are assumed to make a full recovery from their BSI. Life years accrued for survivors were estimated using US national life tables for 2020 [42]. Survivors of BSI were also assumed to fully recover their previous health-related quality of life (HRQoL). Life years accrued by survivors of BSI were weighted using population data on HRQoL to calculate QALYs accrued over their remaining life expectancy. Data on HRQoL for the US population was taken from a multi-national study which summarised data from 20 countries [43]. Time trade-off values were used for the US. The data applied for the US are tabulated below (Table 12). Data on life expectancy for the UK (scenario analysis) were taken from the Office for National Statistics [44]. Quality of life weights for the UK were taken from analysis reported by the National Institute for Health and Care Excellence (NICE) Decision Support Unit [45].

Table 12: HRQoL tariffs as a function of age and sex for the US population.

| **Age-band** | **Males** | **Females** |
| --- | --- | --- |
| 0-24 years | 0.90 | 0.90 |
| 25-34 years | 0.88 | 0.88 |
| 35-44 years | 0.85 | 0.85 |
| 45-54 years | 0.81 | 0.81 |
| 55-64 years | 0.78 | 0.78 |
| 65-74 years | 0.76 | 0.76 |
| 75 years and older | 0.68 | 0.68 |

Abbreviations: HRQoL, health-related quality of life; US, United States.

3.4. Parameter ranges in sensitivity analyses for key parameters

Parameter ranges in sensitivity analysis for key parameters.

| **Parameter** | **Base case** | **Lower limit** | **Upper limit** | **Distribution in PSA** |
| --- | --- | --- | --- | --- |
| Cohort age | 65.1 | 52.1 | 78.1 | Normal |
| Proportion female | 0.45 | 0.360 | 0.540 | Beta |
| Gram-negative pathogen prevalence | 0.333 | 0.267 | 0.400 | Dirichlet |
| Fungal pathogen prevalence | 0.040 | 0.032 | 0.048 | Dirichlet |
| Time to identification with SoC | 24.0 | 14.4 | 33.6 | Gamma |
| Time to identification of pathogen and resistance mechanisms, mRDT | 3.0 | 2.4 | 3.6 | Gamma |
| Time to AST/resistance mechanisms with SoC | 48.0 | 28.8 | 67.2 | Gamma |
| Duration ratio, LOS, each hour of delay to effective treatment | 1.004 | 1.001 | 1.008 | Log-normal |
| LOS in patients receiving effective broad-spectrum treatment (days) | 11.56 | 8.15 | 14.97 | Gamma |
| Additional LOS with *C. difficile* Infection | 3.66 | 0.92 | 6.40 | Gamma |
| Additional LOS with AKI | 4.31 | 3.45 | 5.17 | Gamma |
| Mortality associated with *C. difficile* Infection | 8.60% | 6.40% | 10.90% | Beta |
| Risk of AKI per day | 1.51% | 1.21% | 1.81% | Beta |
| Mortality associated with AKI | 12.90% | 10.32% | 15.48% | Beta |
| Cobas Eplex BCID panels, cost ($) | 115.00 | 92.00 | 138.00 | Not varied |
| BioFire BCID panel, cost ($) | 110.00 | 88.00 | 132.00 | Gamma |
| BioFire BCID2 panel, cost ($) | 115.00 | 92.00 | 138.00 | Gamma |
| Diasorin Verigene, cost ($) | 64.00 | 51.20 | 76.80 | Gamma |
| Accelerate PhenoTest BC kit, cost ($) | 168.00 | 134.40 | 201.60 | Gamma |
| Hospitalisation cost per day ($) | 3025.00 | 2420.00 | 3630.00 | Gamma |
| De-escalation following pathogen ID | 62.9% | 50.3% | 75.5% | Beta |

Abbreviations: AKI, acute kidney injury; BCID, blood culture identification; ID, identification; LOS, length of stay; mRDT, molecular rapid diagnostic test; PSA, probabilistic sensitivity analysis; SoC. standard-of-care.

4. Results

A detailed breakdown of the base case results is presented in Table 13.

Table 13: Breakdown of results in the base case.

| **Diagnostic strategy** | **Overall cost (per patient)** | **Cost saving compared to SoC** | **QALYs gained (per patient** | **QALY gain compared to SoC** | **Deaths (per 10,000 patients)** | **Lives saved compared to SoC (per 10,000 patients)** | **NMB (at $50,000 per QALY)** |
| --- | --- | --- | --- | --- | --- | --- | --- |
| Conventional culture and MALDI-ToF MS (SoC) | $35,515 | ̶ | 8.917 | ̶ | 1,127.8 | ̶ | $410,339 |
| Cobas Eplex BCID panels | $35,351 | $164 | 8.941 | 0.024 | 1,103.6 | 24.2 | $411,719 |
| BioFire BCID panel | $35,372 | $144 | 8.939 | 0.022 | 1,105.9 | 21.9 | $411,585 |
| BioFire BCID2 panel | $35,364 | $151 | 8.940 | 0.023 | 1,104.7 | 23.1 | $411,649 |
| Diasorin Verigene BCID panels | $35,365 | $151 | 8.934 | 0.017 | 1,111.1 | 16.7 | $411,331 |
| Accelerate PhenoTest BC Kit | $35,456 | $59 | 8.937 | 0.020 | 1,108.3 | 19.5 | $411,378 |

Abbreviations: BCID, blood culture identification; MALDI-ToF MS, matrix-assisted laser desorption/ionisation - time of flight mass spectrometry; NMB, net monetary benefit; QALY, quality-adjusted life year; SoC, standard-of-care.

The one-way sensitivity analysis (OWSA) comparing Cobas Eplex BCID panels with SoC (MALDI-ToF MS) is shown in Figure 6. The parameter with the largest impact on the incremental net monetary benefit (INMB) for Cobas Eplex BCID panels is the time taken to identify a pathogen with SoC. The INMB for Cobas Eplex BCID panels remains positive when the time taken to identify a pathogen with MALDI-ToF MS is reduced to 14.4 hours. The second most influential parameter is the cohort age. The impact of varying the remaining parameters is modest.

Figure 6: Tornado plot for the OWSA of Cobas Eplex BCID panels versus SoC (MALDI-ToF MS)

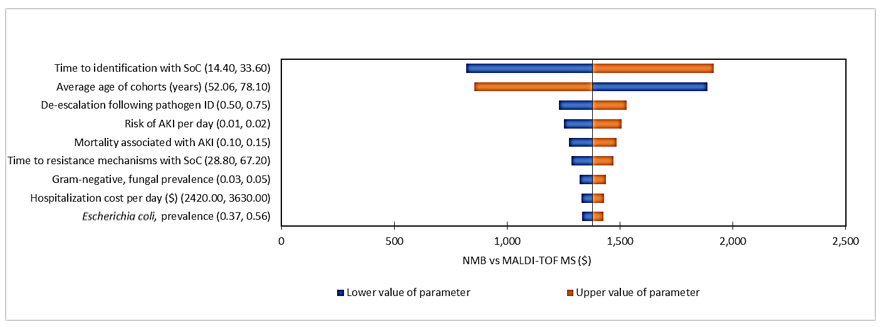

Abbreviations: AKI, acute kidney injury; MALDI-ToF MS, matrix-assisted laser desorption/ionisation - time of flight mass spectrometry; NMB, net monetary benefit; OWSA, one-way sensitivity analysis; SoC, standard-of-care.

4.1. Scenario analyses

The results of the scenario analysis in a UK setting are shown in Table 14. The cheapest comparator is Diasorin Verigene BCID panels. This comparator dominates Accelerate PhenoTest BC kit and SoC. BioFire BCID panel and BioFire BCID2 panel are more effective than Diasorin Verigene BCID panels, but are dominated by Cobas Eplex BCID panels. The incremental cost-effectiveness ratio (ICER) for Cobas Eplex BCID panels compared to Diasorin Verigene BCID panels is £14,190. This means that a QALY is gained for each additional £14,190 spent on testing with Cobas Eplex BCID panels compared to Diasorin Verigene panels. This cost would be considered acceptable in a UK setting, and hence the cost-effective option is Cobas Eplex BCID panels.

Table 14: Scenario analysis in a UK setting

| **Diagnostic strategy** | **Overall cost** | **QALYs** | **Deaths (per 10,000 patients)** | **ICER** |
| --- | --- | --- | --- | --- |
| Conventional culture and MALDI-ToF MS (SoC) | £14,330 | 9.656 | 1,144.7 | Dominated |
| Accelerate PhenoTest BC Kit | £14,386 | 9.673 | 1,129.0 | Dominated |
| Diasorin Verigene panels | £14,241 | 9.677 | 1,125.5 | - |
| BioFire BCID panel | £14,285 | 9.678 | 1,124.3 | Dominated |
| BioFire BCID2 panel | £14,285 | 9.679 | 1,123.5 | Dominated |
| Cobas Eplex BCID panels | £14,280 | 9.680 | 1,122.9 | £14,190 |

Abbreviations: BCID, blood culture identification; ICER, incremental cost-effectiveness ratio; MALDI-ToF, Matrix-assisted laser desorption/ionisation - time of flight; QALY, quality-adjusted life year; SoC, standard-of-care; UK, United Kingdom.

Table 15 and Table 16 presents the results of the scenarios in which genus calls, and both genus and group calls are excluded. Costs rise modestly for mRDT when the value of information on either genus calls, or both group and genus calls are discarded, and QALY gains for mRDT reduce compared with the base case. However, the ranking of comparators remains the same and Cobas Eplex BCID panels remain dominant over all other comparators.

Table 15: Scenario analysis in which genus calls are excluded

| **Diagnostic strategy** | **Overall cost** | **QALYs** | **Deaths (per 10,000 patients)** |
| --- | --- | --- | --- |
| Conventional culture and MALDI-ToF MS (SoC) | $35,515 | 8.917 | 1127.8 |
| Diasorin Verigene panels | $35,421 | 8.930 | 1115.4 |
| Accelerate PhenoTest BC Kit | $35,421 | 8.930 | 1,115.4 |
| BioFire BCID panel | $35,453 | 8.933 | 1,112.3 |
| BioFire BCID2 panel | $35,414 | 8.936 | 1,108.6 |
| Cobas Eplex BCID panels | $35,397 | 8.938 | 1,107.1 |

Abbreviations: BCID, blood culture identification; MALDI-ToF, Matrix-assisted laser desorption/ionisation - time of flight; QALY, quality-adjusted life year; SoC, standard-of-care.

Table 16: Scenario analysis in which both genus and group calls are excluded

| **Diagnostic strategy** | **Overall cost** | **QALYs** | **Deaths (per 10,000 patients)** |
| --- | --- | --- | --- |
| Conventional culture and MALDI-ToF MS (SoC) | $35,515 | 8.917 | 1,127.8 |
| Diasorin Verigene panels | $35,421 | 8.930 | 1,115.4 |
| Accelerate PhenoTest BC Kit | $35,542 | 8.930 | 1,115.0 |
| BioFire BCID panel | $35,453 | 8.933 | 1,112.3 |
| BioFire BCID2 panel | $35,431 | 8.935 | 1,109.9 |
| Cobas Eplex BCID panels | $35,414 | 8.937 | 1,108.5 |

Abbreviations: BCID, blood culture identification; MALDI-ToF, Matrix-assisted laser desorption/ionisation - time of flight; QALY, quality-adjusted life year; SoC, standard-of-care.

In the scenario in which all pathogens detected are considered to be invasive species, costs rise for all comparators including SoC (Table 17). Cost savings for mRDT increase when compared with SoC. In this scenario deaths increase considerably for all comparators and QALYs gained fall; the ranking of comparators remains unchanged from the base case and the Cobas Eplex BCID panels continues to dominate all other comparators.

Table 17: Scenario analysis in which all pathogens detected are considered invasive species

| **Diagnostic strategy** | **Overall cost** | **QALYs** | **Deaths (per 10,000 patients)** |
| --- | --- | --- | --- |
| Conventional culture and MALDI-ToF MS (SoC) | $35,564 | 8.110 | 1,930.6 |
| Diasorin Verigene panels | $35,396 | 8.129 | 1,912.1 |
| Accelerate PhenoTest BC Kit | $35,489 | 8.132 | 1,909.6 |
| BioFire panels | $35,403 | 8.134 | 1,906.9 |
| BioFire BCID2 panel | $35,395 | 8.135 | 1,905.7 |
| Cobas Eplex BCID panels | $35,376 | 8.137 | 1,904.0 |

Abbreviations: BCID, blood culture identification; MALDI-ToF, Matrix-assisted laser desorption/ionisation - time of flight; QALY, quality-adjusted life year; SoC, standard-of-care.

Table 18 reports the scenario analysis examining the proportion of patients for whom empiric therapy is effective whose treatment is optimised to a targeted therapy following pathogen identification. In the scenario this was reduced to 30% from the base case value of 62.9%. In this scenario costs rise for all comparators compared to the base case, but they rise far more sharply for mRDT than for SoC. Despite this, all mRDT remain less costly overall than SoC, and Cobas Eplex BCID panels remain dominant over all other comparators.

Table 18: Scenario analysis in which the proportion of patients optimised to targeted therapy from effective empiric therapy is reduced

| **Diagnostic strategy** | **Overall cost** | **QALYs** | **Deaths (per 10,000 patients)** |
| --- | --- | --- | --- |
| Conventional culture and MALDI-ToF MS (SoC) | $35,705 | 8.917 | 1,127.8 |
| Diasorin Verigene panels | $35,627 | 8.928 | 1,116.5 |
| Accelerate PhenoTest BC kit | $35,712 | 8.932 | 1,113.4 |
| BioFire BCID panel | $35,635 | 8.933 | 1,111.5 |
| BioFire BCID2 panel | $35,630 | 8.934 | 1,110.6 |
| Cobas Eplex BCID panels | $35,622 | 8.935 | 1,109.8 |

Abbreviations: BCID, blood culture identification; MALDI-ToF, Matrix-assisted laser desorption/ionisation - time of flight; QALY, quality-adjusted life year; SoC, standard-of-care.

[7]	"BIOFIRE® Blood Culture Identification 2 (BCID2) Panel. Product information. Available at: <https://www.biomerieux.com/corp/en/our-offer/clinical-products/biofire-blood-culture-identification-2-panel.html>? Accessed 03/02/25.."

[8]	"Verigene® Gram-Negative Blood Culture Nucleic Acid Test (BC-GN). Product information. Available at: <http://www.nanosphere.us/sites/default/files/support-docs/nanosphere_bcgn_insert_final.pdf>. Accessed 03/02/25."

[9]	"VERIGENE Gram-Positive Blood Culture Test. Product information. Available at: <https://int.diasorin.com/en/molecular-diagnostics/kits-reagents/verigene-gram-positive-blood-culture-test-bc-gp>. Accessed 03/02/25.."

[17]	Blood Stream Infection: Focus on Outcome Study Group, "Poster: Results of a Prospective Randomised Multicentre Trial to assess the Impact of Laboratory Based Rapid Diagnostics using MALDI-TOF Technology on Outcomes of Patients with Bood Stream Infection (BSI) (RAPIDO Study)," *ECCMID Vienna,* 2017.

[45]	M. Hernández Alava and S. A. W. Pudney, " Estimating EQ-5D by Age and Sex for the UK. NICE DSU Report," 2022.

Appendices

Appendix A: Escalation examples of ineffective empiric therapy in suspected BSI patients (empiric therapy is too narrow)

| **Pathogen detected** | **Empiric Therapy** | **Ineffectiveness** |
| --- | --- | --- |
| ESBL-producing Enterobacteriaceae | Ceftriaxone or piperacillin-tazobactam | These antibiotics may not cover ESBL-producing E. coli or Klebsiella. Carbapenems are needed. |
| MRSA bloodstream infection | Cefazolin or nafcillin | Ineffective against MRSA; vancomycin or daptomycin is required. |
| VRE bloodstream infection | Vancomycin | Ineffective against VRE; linezolid or daptomycin should be used. |
| Multidrug-resistant Pseudomonas aeruginosa | Piperacillin-tazobactam or cefepime | Ineffective if the strain is resistant; ceftolozane-tazobactam or cefiderocol may be needed. |
| CRE | Meropenem or imipenem | Ineffective if the pathogen is CRE; ceftazidime-avibactam or meropenem-vaborbactam may be required. |
| Candida bloodstream infection | Ceftriaxone or piperacillin-tazobactam | Ineffective as antibiotics don't target fungi; fluconazole or echinocandins (e.g., caspofungin) should be used. |
| Acinetobacter baumannii bloodstream infection | Ceftriaxone or piperacillin-tazobactam | If the strain is multidrug-resistant, therapy may need to escalate to agents like colistin or tigecycline. |

Abbreviations: BSI, bloodstream infection; CRE, carbapenem-resistant Enterobacteriaceae; E. Coli, escherichia coli; ESBL, extended spectrum beta-lactamase; MRSA, methicillin-resistant Staphylococcus aureus; VRE, vancomycin-resistant enterococci.

Appendix B: De-escalation examples of ineffective empiric therapy in suspected BSI (empiric therapy is too broad)

| **Pathogen detected** | **Empiric Therapy** | **Ineffectiveness** |
| --- | --- | --- |
| MSSA bloodstream infection | Vancomycin or daptomycin | Vancomycin is not needed for MSSA and should be de-escalated to cefazolin or nafcillin. |
| Non-ESBL-producing Enterobacteriaceae | Meropenem or piperacillin-tazobactam | Carbapenems are unnecessary if the pathogen is non-ESBL-producing E. coli or Klebsiella; de-escalate to ceftriaxone. |
| CoNS | Vancomycin and piperacillin-tazobactam | CoNS are often contaminants or susceptible to narrower-spectrum antibiotics, so broad-spectrum therapy is often unnecessary. |
| Pseudomonas aeruginosa with low resistance | Meropenem or piperacillin-tazobactam | De-escalation can occur to narrower-spectrum agents (e.g., cefepime) if resistance is not detected. |
| Streptococcus pneumoniae bloodstream infection | Vancomycin and ceftriaxone | Vancomycin can be de-escalated once S. pneumoniae is identified and beta-lactams like ceftriaxone are adequate. |
| Enterobacter spp. susceptible to cefepime | Meropenem or piperacillin-tazobactam | De-escalation to cefepime is recommended if susceptibility testing shows effectiveness. |
| Candida non-albicans species | Echinocandins (e.g., caspofungin) | If the species is susceptible to fluconazole, de-escalation from echinocandins is appropriate. |

Abbreviations: BSI, bloodstream infection; CoNS, coagulase-negative staphylococci; E. coli; escherichia coli; ESBL, extended spectrum beta-lactamase; MSSA, meticillin-sensitive staphylococcus aureus; spp, several species.

Appendix C: Examples of stopping empiric antibiotic therapy

| **Pathogen detected** | **Empiric Therapy** | **Ineffectiveness** |
| --- | --- | --- |
| Viral bloodstream infection (e.g., Influenza, COVID-19) | Broad-spectrum antibiotics (e.g., ceftriaxone, vancomycin) | If PCR or serologic testing confirms a viral pathogen, such as influenza or SARS-CoV-2, antibiotics should be stopped, as they do not treat viral infections. |
| Positive blood cultures for CoNS | Vancomycin or ceftriaxone | CoNS are common blood culture contaminants, especially in patients with indwelling devices. If cultures are identified as contaminants and the patient has no signs of sepsis, antibiotics can be stopped. |
| Negative blood cultures after 48-72 hours | Broad-spectrum antibiotics (e.g., piperacillin-tazobactam, cefepime) | If blood cultures remain negative after 48-72 hours and there is no clinical evidence of bacterial infection, antibiotics should be discontinued. |
| Contaminated blood cultures with skin flora (e.g., Corynebacterium, Bacillus spp.) | Vancomycin or ceftriaxone | Certain organisms (e.g., Corynebacterium, Bacillus spp.) are often contaminants from skin flora. If determined to be contaminants, antibiotics can be stopped. |
| Non-infectious cause of fever (e.g., drug fever, autoimmune disease) | Broad-spectrum antibiotics (e.g., meropenem, vancomycin) | If fever or symptoms are determined to be caused by non-infectious factors (e.g., drug reaction or autoimmune flare), antibiotics can be discontinued. |
| Confirmed fungal or parasitic infection | Empiric antibiotics (e.g., ceftriaxone, piperacillin-tazobactam) | If a fungal or parasitic infection is confirmed (e.g., Candida or Plasmodium), antibiotics should be stopped and appropriate antifungal or antiparasitic treatment initiated. |
| Proven diagnosis of DVT or PE | Empiric antibiotics (e.g., ceftriaxone, vancomycin) | If a patient’s fever or inflammatory response is due to a clotting disorder like DVT or PE, and not infection, antibiotics can be stopped. |
| Suspected bacterial infection but no clinical or microbiological evidence | Broad-spectrum antibiotics (e.g., piperacillin-tazobactam) | If diagnostic tests (e.g., imaging, cultures) and clinical follow-up do not support an infectious etiology, antibiotics should be stopped. |
| Unexplained fever resolved after other treatment | Broad-spectrum antibiotics (e.g., ceftriaxone, vancomycin) | If the fever resolves without evidence of infection (e.g., after antipyretics or treatment of another underlying condition), empiric antibiotics should be stopped. |
| Post-surgical fever without infection | Broad-spectrum antibiotics (e.g., ceftriaxone, vancomycin) | Post-operative fevers are common and often not caused by infection. If no signs of infection are found, antibiotics can be stopped. |

Abbreviations: CoNS, coagulase-negative staphylococci; DVT, deep vein thrombosis; PCR, polymerase chain reaction; PE, pulmonary embolism; spp, several species.
