## Supplementary material for "Evaluating the cost-effectiveness of rapid diagnostic testing for the identification of pathogens and resistance genes in bloodstream infections": CHEERS statement

**The Consolidated Health Economic Evaluation Reporting Standards (CHEERS) checklist - 2022**

1. **Title**

Identify the study as an economic evaluation and specify the interventions being compared.

Location where item is reported: Title

2. **Abstract**

Provide a structured summary that highlights context, key methods, results, and alternative analyses.

Location where item is reported: structured abstract

3. **Background and objectives**

Give the context for the study, the study question, and its practical relevance for decision making in policy or practice.

Location where item is reported: introduction

4. **Health economic analysis plan**

Indicate whether a health economic analysis plan was developed and where available.

Location where item is reported: Supplementary material, section S1.1

5. **Study population**

Describe characteristics of the study population (such as age range, demographics, socioeconomic, or clinical characteristics).

Location where item is reported: Methods, section 2.1, first paragraph

6. **Setting and location**

Provide relevant contextual information that may influence findings.

Location where item is reported: Methods, section 2.1

7. **Comparators**

Describe the interventions or strategies being compared and why chosen.

Location where item is reported: Methods S2.2

8. **Perspective**

State the perspective(s) adopted by the study and why chosen.

Location where item is reported: Methods, section 2.1

9. **Time horizon**

State the time horizon for the study and why appropriate.

Location where item is reported: Methods, section 2.1, first paragraph

10. **Discount rate**

Report the discount rate(s) and reason chosen.

Location where item is reported: Methods, section 2.1, first paragraph

11. **Selection of outcomes**

Describe what outcomes were used as the measure(s) of benefit(s) and harm(s).

Location where item is reported: Methods, section 2.1, first paragraph

12. **Measurement of outcomes**

Describe how outcomes used to capture benefit(s) and harm(s) were measured.

Location where item is reported: Methods, section 2.1, third paragraph

13. **Valuation of outcomes**

Describe the population and methods used to measure and value outcomes.

Location where item is reported: Methods, section 2.1, fourth paragraph

14. **Measurement and valuation of resources and costs**

Describe how costs were valued.

Location where item is reported: Methods, section 2.3, paragraphs eight and nine

15. **Currency, price date, and conversion**

Report the dates of the estimated resource quantities and unit costs, plus the currency and year of conversion.

Location where item is reported: Methods, section 2.3, paragraphs nine

16. **Rationale and description of model**

If modelling is used, describe in detail and why used. Report if the model is publicly available and where it can be accessed.

Location where item is reported: Methods, section 2.1

17. **Analytics and assumptions**

Describe any methods for analysing or statistically transforming data, any extrapolation methods, and approaches for validating any model used.

Location where item is reported: Methods, section 2.3 and supplementary materials section 3.

18. **Characterising heterogeneity**

Describe any methods used for estimating how the results of the study vary for subgroups.

Location where item is reported: N/A

19. **Characterising distributional effects**

Describe how impacts are distributed across different individuals or adjustments made to reflect priority populations.

Location where item is reported: N/A

20. **Characterising uncertainty**

Describe methods to characterise any sources of uncertainty in the analysis.

Location where item is reported: Methods, section 2.4

21. **Approach to engagement with patients and others affected by the study**

Describe any approaches to engage patients or service recipients, the general public, communities, or stakeholders (such as clinicians or payers) in the design of the study.

Location where item is reported: Supplementary materials: section 1.1

22. **Study parameters**

Report all analytic inputs (such as values, ranges, references) including uncertainty or distributional assumptions.

Location where item is reported: Methods, section 2.3

23. **Summary of main results**

Report the mean values for the main categories of costs and outcomes of interest and summarise them in the most appropriate overall measure.

Location where item is reported: Results, section 3.1

24. **Effect of uncertainty**

Describe how uncertainty about analytic judgments, inputs, or projections affect findings. Report the effect of choice of discount rate and time horizon, if applicable.

Location where item is reported: Results, section 3.2

25. **Effect of engagement with patients and others affected by the study**

Report on any difference patient/service recipient, general public, community, or stakeholder involvement made to the approach or findings of the study

Location where item is reported: N/A

26. **Study findings, limitations, generalisability, and current knowledge**

Report key findings, limitations, ethical or equity considerations not captured, and how these could affect patients, policy, or practice.

Location where item is reported: Discussion, sections 4.1 and 4.2

**Other relevant information**

27. **Source of funding**

Describe how the study was funded and any role of the funder in the identification, design, conduct, and reporting of the analysis

Location where item is reported: Declaration of funding statement with abstract

28. **Conflicts of interest**

Report authors conflicts of interest according to journal or International Committee of Medical Journal Editors requirements.

Location where item is reported: conflict of interest statement with abstract
